# Prevalence and Predictors of Cigarette Smoking Among School-Going Adolescents in Africa Based on the Global Youth Tobacco Survey: 2001-2021

**DOI:** 10.64898/2026.02.19.26346679

**Authors:** Jerry Sinyangwe, Wingston Felix Ng’ambi, Cosmas Zyambo

**Affiliations:** Department of Community and Family Medicine, University of Zambia, Lusaka, Zambia; Health Economics and Policy Unit, Department of Health Systems and Policy, Kamuzu University of Health Sciences, Lilongwe, Malawi

**Keywords:** Adolescent tobacco use, cigarette smoking, Africa, Global Youth Tobacco Survey, tobacco control

## Abstract

**Introduction:** Tobacco smoking among school-going adolescents poses a major global public health challenge, contributing significantly to future disease burden. Understanding the prevalence and determinants of cigarette smoking in this group is critical for effective interventions.

**Methods:** This study analyzed data from the Global Youth Tobacco Surveys collected between 2001 and 2021, covering 439, 322 school-going adolescents aged 11 to 19 across 45 African countries. Descriptive statistics estimated smoking prevalence by age, sex, school grade, country, and survey year. Predictive modelling identified independent correlates of current cigarette smoking under complex two-stage cluster sampling.

**Results:** Overall, 23.8% (95% CI: 20.20-27.57) of adolescents reported cigarette smoking. Prevalence increased with age, rising from 9.5% at 12 years to 22.4% at 18 years. Boys smoked more than girls (14.0% vs. 7.3%). Smoking varied widely across countries, with the highest rates in Burkina Faso 48.9%, and South Africa (18.9%), and the lowest in Angola (1.5%, 95% CI: 0.86-2.13), Eritrea (2.1%). Use of other tobacco products strongly increased the prevalence of cigarette smoking for smokeless chew (35%, 95% CI: 29.6–40.3). Being taught in school about the effects of smoking showed protective effects, while ownership of tobacco-branded items increased smoking likelihood (17.8%, 95% CI: 17.52-18.13). Smoking prevalence declined over time, with lowest rates in recent years (3.5% in 2020).

**Conclusion:** Cigarette smoking among school-going adolescents in Africa is a growing public health concern influenced by factors such as age, gender, country, behavior, and media exposure. Urgent, youth-focused tobacco control strategies especially targeting males and older teens are needed. Strengthening school-based education and implementing tobacco control policies among students can help reduce smoking rates. These findings offer vital evidence to inform global tobacco control efforts within the African context

## INTRODUCTION

Tobacco smoking remains one of the leading preventable causes of illness and premature death worldwide (1,2). Globally, over 1.3 billion people use tobacco products, with cigarette smoking accounting for the majority of tobacco-related morbidity and mortality (3,4). Despite decades of tobacco control efforts, the World Health Organization (WHO) estimates that tobacco use kills more than 8 million people annually (5), including around 1.2 million deaths caused by secondhand smoke exposure (6). Adolescence is a critical period when tobacco habits are often formed, with early initiation strongly linked to lifelong addiction and increased risk of non-communicable diseases such as cardiovascular disease, cancer, and chronic respiratory conditions (7,8).

In Africa, cigarette smoking patterns reflect a complex interplay of cultural, social, and economic factors (9), along with varying enforcement of tobacco control policies (8,10). While overall smoking prevalence remains lower in many African countries (11,12) compared to global averages, recent trends indicate rising tobacco use among youths, especially in urban settings (13). The Global Youth Tobacco Survey (GYTS) and other regional studies show that approximately 10% of African school-going adolescents currently smoke cigarettes with significant variations by age, sex, and country (14,15). Male adolescents consistently report higher smoking rates than females, reflecting gender norms and marketing strategies targeting young men (14,16,17). Meanwhile, exposure to tobacco advertising (18) and weak enforcement of tobacco advertising and promotion bans continue to hinder efforts to reduce youth tobacco use (19,20).

This study will analyze secondary data from the GYTS in 45 African countries. Adolescents are a crucial group for monitoring tobacco use since experimentation and initiation often start during this developmental stage (21). Most smokers start young (22) with a significant number of adult smokers having started before the age of 25 years (23). Early onset tobacco use is harmful to adolescents whose brains are still developing affecting attention, learning and impulse control (22). Early tobacco use also predicts lifelong addiction (21). School going adolescents are targeted for collecting data during the surveys because they are easily accessible and the school environment enhances standardized sampling and data comparability across countries (24). Using schools as survey sites enhances logistical feasibility and facilitates efficient data collection (24).

This study aims to assess the prevalence and determinants of current cigarette smoking among school-going adolescents across Africa. It focuses on how demographic, behavioral, and policy-related factors influence smoking behaviors over time. Specifically, the study estimates smoking prevalence among youths aged 11 to 19 years in 45 African countries, exploring variations by age, sex, school grade, and country. It identifies key correlates of smoking including the use of other tobacco products and exposure to tobacco advertising and anti-smoking media. Together, these findings will provide valuable evidence to guide policy makers, educators, and public health practitioners in designing effective tobacco control strategies tailored to African youth populations. This study aligns with the global tobacco research agenda by advancing understanding of tobacco use dynamics in low- and middle-income countries, where data have historically been limited but are crucial for global progress toward tobacco control targets.

## METHODS

### Data Source

This study used data from the Global Youth Tobacco Survey (GYTS), collected between 2001 and 2021 across 45 African countries. The survey targeted school-going adolescents aged 11 to 19 years, gathering detailed information on tobacco use, exposure to tobacco advertising, anti-smoking media awareness, school-based tobacco education, and perceptions of tobacco control policies. The GYTS is designed to generate a cross sectional national representative sample. It has a standardized methodology for the sampling framework, selection of schools and classes and data processing. The survey employs a standard questionnaire with core questions and optional questions which allow adaptation to the needs of a particular country. A more detailed description of the overall objectives and methodology of the GYTS are available elsewhere (24). The schools and sample size are determined using a standard procedure and software developed by the Center for Disease Control and Prevention (CDC) once it receives the sampling frame from the GYTS research coordinators. The actual sample size and its computation was not available, however, a minimum sample size of 1500 students, 50% of whom are female is required for the GYTS (24). A total of 439,322 school going adolescents from 45 countries in Africa were surveyed between 2001 and 2021. Each country’s survey used a two-stage cluster sampling design to ensure nationally representative samples of school going youths.

### Data management

The primary outcome variable was current cigarette smoking that was generated from the question, “During the past 30 days (one month), on how many days did you smoke cigarettes (including manufactured or hand-rolled cigarettes)?” with response options: 0, 1 or 2, 3 to 5, 6 to 9, 10 to 19, 20 to 29, all 30 days. Participants who smoked cigarettes at least once (Yes) in the last 30 days were classified as current cigarette smokers, while those who did not (No) were classified as non-current cigarette smokers.

### Independent variables

#### Socio-demographic factors

These were: age (≥11, 12, 13, 14, 15, 16, 17, 18, and ≤ 19 years), sex (male and female) and grade (6, 7, 8, 9, 10, 11, 12, 13 and ≤ 14).

#### Behavioral and tobacco marketing factors

Our study used seven questions from the questionnaire to which the students answered “Yes” or “No”: 1) “During the last 30 days, did you ever try smokeless chewed tobacco?” 2) “During the last 30 days, did you ever try using snuff?” 3) “During the last 30 days, did you ever try smoking shisha?” 4) “During the last 30 days, did you ever use chewing tobacco?” Students who answered “Yes” were classified as smokeless chewed tobacco, snuff, shisha and chewing tobacco users respectively otherwise they were not. Participants were also asked; 5) “During the past 30 days, how often did you see any people using tobacco products on TV, the internet or social media (Facebook, Twitter, Instagram, TikTok or YouTube)?”; 6) “Do you own anything other than a tobacco product (for example, t-shirt, pen, backpack, hat, and ball) with a tobacco product brand logo on it?” Students who answered “Yes” in either case were classified as having been exposed to tobacco marketing; Students who responded “Yes” to the question, 6) “During the past 30 days, did you see or hear any messages against the use of tobacco products in the media?” were classified as having been exposed to anti-tobacco use adverts in the media otherwise they were not.

### School Curriculum factors

This was generated from the question, “During the past 12 months, were you taught in classes at school about the dangers of using tobacco products?” To which the students responded Yes/No.

#### Policy factors

This covariate had four questions to which the students responded Yes/No; 1) “Do you support banning of tobacco smoking in public?”; 2) “Do you support banning of tobacco sales to minors (under 18)?”; 3) “Do you support banning of tobacco advertising and promotions?”; 4) “Are you aware of the tobacco act?”

Categorical variables were recoded into binary or grouped factors as appropriate to enhance clarity and consistency. Missing data were addressed using imputation techniques based on observed distributions or logical carry-forward from related variables, aimed at reducing bias and improve completeness. For example, missing smoking status was imputed using probabilities derived from respondents with similar characteristics.

#### Data Cleaning and Preprocessing

We ensured consistent coding across different countries and years by standardizing variable labels and responses. Age values missing, less than 11 years or above 19 years were replaced with the median (15 years) or capped at 19. Missing sex data were imputed according to observed gender proportions. Missing school grades or exceeding 14 were set to the mode (grade 9) or capped at 14. All categorical data were converted to categorical variables for analysis, and clear labels were assigned for ease of interpretation. A composite indicator for current smoking was created using all available tobacco use measures to strengthen the reliability of the outcome.

### Statistical Analysis

Descriptive summaries were applied to obtain frequencies and proportions of current cigarette smokers and non-cigarette smokers across socio-demographic, behavioral, and policy related factors (25). The Chi-square test was used to determine associations between various factors and current cigarette smoking status. Unadjusted logistic regression models were utilized to explore crude relationships between each factor and current cigarette smoking (26). Variables that were found to be significantly associated (p<0.05) with current cigarette smoking status in the univariate analyses were included in the multivariate models, refined by stepwise selection guided by the Akaike Information (AIC) Criterion to identify independent predictors. Multicollinearity assessment using Variance Inflation Factor (IVF), Goodness of fit evaluation (Hosmer-Lemeshow test) and Residual and influence diagnostics were employed as model diagnostics. To complement inferential statistics, Elastic Net Logistic Regression (α=0.5) and Random Forest Classifier machine learning models were used. Machine learning model performance was evaluated using the Area Under the Receiver Operating Characteristic Curve (AUC) and Classification accuracy. All analyses were conducted in R (version 4.3.1), using packages such as tidyverse, gtsummary/gt, glmnet, randomForest and pROC. For descriptive tables (Table 1), broom and MASS for modeling.

**Table 1:** Descriptive Statistics of Study Participants in 45 African countries: 2001-2021.

| Characteristics (n=439,322) | n (%) |
| --- | --- |
| **Country** |  |
| Algeria | 13714 (3.1%) |
| Angola | 1576 (0.4%) |
| Benin | 4329 (1.0%) |
| Botswana | 4127 (0.9%) |
| Burkina Faso | 11081 (2.5%) |
| Burundi | 2521 (0.6%) |
| Cameroon | 15599 (3.6%) |
| Cape Verde | 2019 (0.5%) |
| Central African Republic | 2027 (0.5%) |
| Chad | 5333 (1.2%) |
| Comoros | 4620 (1.1%) |
| Congo | 3109 (0.7%) |
| Democratic Republic of Congo | 12717 (2.9%) |
| Equatorial Guinea | 3136 (0.7%) |
| Eritrea | 9639 (2.2%) |
| Ethiopia | 1868 (0.4%) |
| Gabon | 1781 (0.4%) |
| Gambia | 14544 (3.3%) |
| Ghana | 23949 (5.5%) |
| Guinea | 5038 (1.1%) |
| Ivory Coast | 10152 (2.3%) |
| Kenya | 17411 (4.0%) |
| Lesotho | 7573 (1.7%) |
| Liberia | 1739 (0.4%) |
| Madagascar | 4911 (1.1%) |
| Malawi | 7617 (1.7%) |
| Mali | 6227 (1.4%) |
| Mauritania | 15740 (3.6%) |
| Mauritius | 7812 (1.8%) |
| Mozambique | 13642 (3.1%) |
| Namibia | 8642 (2.0%) |
| Niger | 6193 (1.4%) |
| Nigeria | 5459 (1.2%) |
| Rwanda | 2284 (0.5%) |
| Senegal | 13208 (3.0%) |
| Seychelles | 5314 (1.2%) |
| Sierra Leone | 9611 (2.2%) |
| South Africa | 28370 (6.5%) |
| Sao Tome Principe | 8525 (1.9%) |
| Swaziland | 27211 (6.2%) |
| Tanzania | 16016 (3.6%) |
| Togo | 17879 (4.1%) |
| Uganda | 17452 (4.0%) |
| Zambia | 22263 (5.1%) |
| Zimbabwe | 15344 (3.5%) |
| **Year** |  |
| 2001 | 22626 (5.2%) |
| 2002 | 41637 (9.5%) |
| 2003 | 26756 (6.1%) |
| 2004 | 6231 (1.4%) |
| 2005 | 23145 (5.3%) |
| 2006 | 32930 (7.5%) |
| 2007 | 45376 (10.3%) |
| 2008 | 80144 (18.2%) |
| 2009 | 29087 (6.6%) |
| 2010 | 10101 (2.3%) |
| 2011 | 17660 (4.0%) |
| 2013 | 20748 (4.7%) |
| 2014 | 11130 (2.5%) |
| 2015 | 5295 (1.2%) |
| 2016 | 7981 (1.8%) |
| 2017 | 24929 (5.7%) |
| 2018 | 10118 (2.3%) |
| 2019 | 12609 (2.9%) |
| 2020 | 4320 (1.0%) |
| 2021 | 6499 (1.5%) |
| **Age (years)** |  |
| 11 | 23725 (5.4%) |
| 12 | 39147 (8.9%) |
| 13 | 66148 (15.1%) |
| 14 | 87990 (20.0%) |
| 15 | 88412 (20.1%) |
| 16 | 70843 (16.1%) |
| 17 | 57151 (13.0%) |
| 18 | 5767 (1.3%) |
| 19 | 139 (0.0%) |
| **Sex** |  |
| Female | 220964 (50.3%) |
| Male | 218358 (49.7%) |
| **Grade** |  |
| 6 | 17738 (4.0%) |
| 7 | 59676 (13.6%) |
| 8 | 80567 (18.3%) |
| 9 | 94407 (21.5%) |
| 10 | 85954 (19.6%) |
| 11 | 55764 (12.7%) |
| 12 | 43798 (10.0%) |
| 13 | 1360 (0.3%) |
| 14 | 58 (0.0%) |
| **Used Chewing Tobacco** |  |
| No | 424,650 (96.7%) |
| Yes | 14,672 (3.3%) |
| **Exposed to Smoking Advertising** |  |
| No | 6,092 (1.4%) |
| Yes | 433,230 (98.6%) |
| **Exposed to Anti-Smoking in the Media** |  |
| No | 93,865 (21.4%) |
| Yes | 345,457 (78.6%) |
| **Supported banning of tobacco sales to minors** |  |
| No | 423,951 (96.5%) |
| Yes | 15,371 (3.5%) |
| **Supported banning of tobacco smoking in public** |  |
| No | 177,687 (40.5%) |
| Yes | 261,635 (59.6%) |
| **Supported banning of tobacco advertising** |  |
| No | 431,901 (98.3%) |
| Yes | 7,421 (1.6%) |
| **Owned tobacco branded item** |  |
| No | 370,692 (84.3%) |
| Yes | 68,630 (15.7%) |

## RESULTS

### Characteristics of study participants

This study analysed data from 439,322 school-going youths aged 11 to 19 years. The data covered 45 countries across Africa with South Africa contributing 6.5% while Angola, Gabon, and Liberia, contributed a total of 1.2%. This distribution ensured diverse regional representation, though with varying sample sizes across countries. The sample had a balanced gender distribution, with 50.3% female and 49.7% male (**Table 1**). The majority of the respondents were aged between 13 and 16 years, peaking at age 14 (20.0%) and 15 (20.1%) before declining to 16% and 13% among 16 and 17 year olds respectively. Adolescents aged 11 and 12 years accounted for a combined 14.3% while those aged 18 and 19 years constituted 1.3%. As shown in **Table 1**, adolescents in grades 7-11 accounted for the majority of the respondents, especially those in grades nine (21.5%) and ten (19.6%).

Almost all the students (98.6%) had seen smoking advertisements and notably 15.7% owned a tobacco branded item, while 78.6% reported exposure to anti-smoking messages. A small number of participants reported having used chewing tobacco (3.3%).

Support for banning of tobacco smoking in public was 59.6%, while that of tobacco sales to minors (3.5%) and tobacco advertising (1.6%) were surprisingly low.

### Prevalence of cigarette smoking

**Table 2** shows the prevalence of cigarette smoking across demographic, behavioral, environmental, and policy-related factors among students included in the study (n = 439, 322). Some of the highest cigarette smoking rates were found in Burkina Faso (48.93%, 95% CI: 47.96-49.89), South Africa (18.94%, 95% CI: 18.47-19.42), Seychelles (18.84%, 95% CI: 17.75-19.94), and Mali (18.01%, 95% CI: 17.04-18.98). In contrast, low rates were observed in Angola (1.50%, 95% CI: 0.86-2.13), Eritrea (2.07%, 95% CI: 1.78-2.36) and Liberia (2.60%, 95% CI: 1.82-3.38). Smoking prevalence was highest in 2006 (20.85%, 95% CI: 20.40-21.30) and 2011 (17.70%, 95% CI: 17.12-18.28), and the lowest were recorded in 2019 (3.86%, 95% CI: 3.51-4.21) and 2020 (3.52%, 95% CI: 2.95-4.08).

**Table 2:** Prevalence of cigarette smoking in 45 African countries: 2001-2021.

| Characteristics | Prevalence (%) | Lower 95% CI (%) | Upper 95% CI (%) |
| --- | --- | --- | --- |
| **Country** | 6.00 | 5.60 | 6.40 |
| Angola | 1.33 | 0.77 | 1.90 |
| Benin | 10.30 | 9.40 | 11.21 |
| Botswana | 11.46 | 10.49 | 12.43 |
| Burkina faso | 97.75 | 97.32 | 98.19 |
| Burkina Faso | 10.79 | 10.05 | 11.54 |
| Burundi | 5.24 | 4.37 | 6.11 |
| Cameroon | 6.51 | 6.12 | 6.89 |
| Cape Verde | 5.30 | 4.32 | 6.28 |
| CAR | 7.94 | 6.77 | 9.12 |
| Chad | 6.39 | 5.74 | 7.05 |
| Comoros | 5.17 | 4.53 | 5.81 |
| Congo | 16.95 | 15.63 | 18.27 |
| DRC | 6.97 | 6.52 | 7.41 |
| Equatorial Guinea | 9.57 | 8.54 | 10.60 |
| Eritrea | 1.99 | 1.71 | 2.27 |
| Ethiopia | 2.89 | 2.13 | 3.65 |
| Gabon | 8.65 | 7.34 | 9.95 |
| Gambia | 7.31 | 6.89 | 7.73 |
| Ghana | 4.18 | 3.93 | 4.43 |
| Guinea | 8.63 | 7.86 | 9.41 |
| Ivory Coast | 14.60 | 13.91 | 15.28 |
| Kenya | 7.18 | 6.80 | 7.56 |
| Lesotho | 12.94 | 12.18 | 13.70 |
| Liberia | 2.42 | 1.69 | 3.14 |
| Madagascar | 16.07 | 15.04 | 17.09 |
| Malawi | 2.95 | 2.57 | 3.33 |
| Mali | 17.39 | 16.45 | 18.33 |
| Mauritania | 16.19 | 15.61 | 16.76 |
| Mauritius | 13.04 | 12.30 | 13.79 |
| Mozambique | 2.69 | 2.42 | 2.96 |
| Namibia | 15.04 | 14.29 | 15.80 |
| Niger | 11.38 | 10.59 | 12.17 |
| Nigeria | 4.47 | 3.92 | 5.02 |
| Rwanda | 2.76 | 2.09 | 3.43 |
| Senegal | 9.35 | 8.85 | 9.85 |
| Seychelles | 17.41 | 16.39 | 18.43 |
| Siera Leone | 7.92 | 6.94 | 8.89 |
| Sierraleone | 3.37 | 2.94 | 3.80 |
| South Africa | 17.44 | 17.00 | 17.89 |
| STP | 6.22 | 5.70 | 6.73 |
| Swaziland | 8.62 | 8.28 | 8.95 |
| Tanzania | 2.33 | 2.10 | 2.56 |
| Togo | 6.54 | 6.18 | 6.90 |
| Uganda | 8.39 | 7.98 | 8.81 |
| Zambia | 14.53 | 14.06 | 14.99 |
| Zimbabwe | 5.99 | 5.61 | 6.36 |
| **Year** | 14.04 | 13.59 | 14.49 |
| 2002 | 13.50 | 13.17 | 13.83 |
| 2003 | 8.56 | 8.22 | 8.89 |
| 2004 | 17.01 | 16.08 | 17.94 |
| 2005 | 7.31 | 6.97 | 7.64 |
| 2006 | 19.45 | 19.02 | 19.87 |
| 2007 | 9.22 | 8.95 | 9.49 |
| 2008 | 8.77 | 8.58 | 8.97 |
| 2009 | 8.55 | 8.23 | 8.87 |
| 2010 | 5.45 | 5.01 | 5.90 |
| 2011 | 16.82 | 16.27 | 17.38 |
| 2013 | 3.00 | 2.77 | 3.23 |
| 2014 | 7.44 | 6.95 | 7.93 |
| 2015 | 6.53 | 5.87 | 7.20 |
| 2016 | 7.49 | 6.92 | 8.07 |
| 2017 | 5.09 | 4.82 | 5.37 |
| 2018 | 9.27 | 8.71 | 9.84 |
| 2019 | 3.62 | 3.29 | 3.94 |
| 2020 | 3.31 | 2.78 | 3.84 |
| 2021 | 7.11 | 6.48 | 7.73 |
| **Age** | 10.43 | 10.04 | 10.83 |
| 12 | 8.60 | 8.32 | 8.88 |
| 13 | 7.29 | 7.09 | 7.49 |
| 14 | 8.01 | 7.83 | 8.19 |
| 15 | 9.30 | 9.12 | 9.48 |
| 16 | 11.44 | 11.20 | 11.68 |
| 17 | 13.98 | 13.70 | 14.27 |
| 18 | 19.74 | 18.71 | 20.77 |
| **Sex** | 6.94 | 6.83 | 7.04 |
| Male | 12.73 | 12.59 | 12.87 |
| **Grade** | 9.52 | 9.09 | 9.96 |
| 7 | 7.79 | 7.57 | 8.00 |
| 8 | 9.66 | 9.45 | 9.86 |
| 9 | 8.71 | 8.53 | 8.88 |
| 10 | 10.18 | 9.98 | 10.38 |
| 11 | 11.79 | 11.52 | 12.06 |
| 12 | 12.28 | 11.98 | 12.59 |
| **Chewing tobacco use** | 9.38 | 9.29 | 9.47 |
| Yes | 22.55 | 21.88 | 23.23 |
| **Exposed to smoking advertising** | 6.04 | 5.44 | 6.64 |
| Yes | 9.87 | 9.78 | 9.96 |
| **Exposed to anti-smoking media** | 5.88 | 5.73 | 6.03 |
| Yes | 10.89 | 10.78 | 10.99 |
| **Experimented with cigars** | 9.81 | 9.72 | 9.90 |
| Yes | 14.12 | 11.81 | 16.43 |
| **Tried smokeless chewing tobacco** | 9.80 | 9.71 | 9.89 |
| Yes | 33.75 | 28.57 | 38.93 |
| **Tried smokeless snuff** | 9.81 | 9.72 | 9.89 |
| Yes | 17.33 | 14.48 | 20.17 |
| **Tried shisha/hookah** | 9.78 | 9.69 | 9.87 |
| Yes | 19.61 | 17.74 | 21.48 |
| **School taught tobacco effects** | 10.26 | 9.40 | 11.12 |
| Yes | 9.81 | 9.72 | 9.90 |
| **Tobacco control activities** | 9.82 | 9.73 | 9.91 |
| **Smoking ban for minors** | 9.98 | 9.89 | 10.07 |
| Yes | 5.28 | 4.92 | 5.63 |
| **Public smoking ban** | 11.02 | 10.86 | 11.18 |
| Yes | 9.18 | 9.07 | 9.28 |
| **Tobacco advertising ban** | 9.94 | 9.85 | 10.03 |
| Yes | 2.90 | 2.52 | 3.28 |
| **Owns tobacco-branded item** | 8.72 | 8.63 | 8.81 |
| Yes | 15.75 | 15.48 | 16.02 |

The overall prevalence of cigarette smoking among the respondents was 23.88% (95% CI: 20.20-27.57). The prevalence of current cigarette smoking was highest among 18-year old (22.41%, 95% CI: 21.26-23.57) followed by adolescents aged 17 years (15.29%, 95% CI: 14.98-15.60). Lower prevalence was reported among those aged 19 years (6.98%, 95% CI: 2.58-11.37) and 13 years (8.01%, 95% CI: 7.79-8.23). Cigarette smoking was higher among male (14.01%, 95% CI: 13.86-14.17) than female (7.30%, 95% CI: 7.18-7.41) respondents. By education level, smoking was highest among grades 13 (22.82%, 95% CI: 20.48-25.16) followed by grades 12 (13.39%, 95% CI: 13.05-13.73) and 11 (12.86%, 95% CI: 11.12-11.57).

Behavioral factors related to smoking were equally outlined in **Table 2**. Students who used chewing tobacco had a much higher cigarette smoking prevalence (25.09%, 95% CI: 24.35-25.83) than those who did not (10.33%, 95% CI: 10.23-10.42). Similarly, those who had tried smokeless products like chewed tobacco (34.95%, 95% CI: 29.63-40.27), snuff (21.38%, 95% CI: 17.96-24.80), or shisha (23.50%, 95% CI: 21.31-25.68) reported higher cigarette smoking.

Environmental exposure and policy-related indicators further exemplified tobacco use among adolescents (**Table 2**). Exposure to anti-smoking in the media paradoxically had higher rates (11.79% vs 6.90%). Students who supported bans on smoking in public places had a lower prevalence (10.29%) compared to those who did not (11.64%). Ownership of branded tobacco items was linked to notably higher smoking rates (17.82% vs 9.56%). Interestingly, students who supported banning of tobacco sales to minors or tobacco advertising actually showed lower smoking prevalence. Adolescents who were taught in school about the effects of tobacco use (10.81%, 95% CI: 10.71-10.90) reported a slightly lower smoking prevalence than those who were not taught (11.72%, 95% CI: 10.74-12.70). Students who were aware of the tobacco control act reported lower cigarette smoking as compared to those who were not (5.19% vs. 10.83%).

### Model performance for Elastic Net (α=0.5) and the Random Forest

In this study, two predictive models, Elastic Net (α=0.5) and the Random Forest models were used to correctly rank cigarette smoking and non-smoking cases (Table 3). Both models were comparable in separating classes (AUC 0.762). However, the Random Forest exhibited better accuracy though marginal and therefore provided better prediction reliability.

**Table 3:** Predictive Model Performance Comparison.

| Model | AUC | Accuracy |
| --- | --- | --- |
| Elastic Net ($\alpha=0.5$) | 0.762 | 0.899 |
| Random Forest | 0.762 | 0.907 |

### Predictive probabilities of cigarette smoking

#### Smoking probabilities by country

Figure 1 illustrates the mean predicted cigarette smoking probability by country. The graph reviews distinct variations in the mean predicted probability of smoking in 45 African countries, with the majority showing relatively low predicted probabilities, an indication of a lower estimated risk of cigarette smoking among adolescents. However, a number of countries such as Burkina Faso, Seychelles, South Africa and Mali (Figure 1) had higher values. These differences in predicted probabilities may reflect variations in social norms, tobacco control policies, economic conditions, exposure to tobacco marketing and tobacco product accessibility. The two predictive approaches employed in the analysis produced comparable estimates with minimal differences and country rankings which support the reliability of the observed patterns. The mean predicted probability was marginally higher using the Elastic Net than the Random Forest. For instance, the predictive probabilities for smoking in Burkina Faso using the Elastic Net and the Random Forest were approximately 0.52 and 0.47 respectively. Generally, the results illustrate differences across countries and stress the need to contextualize and employ targeted tobacco prevention and control measures as interventions may yield different results depending on the context.

**Figure 1:**
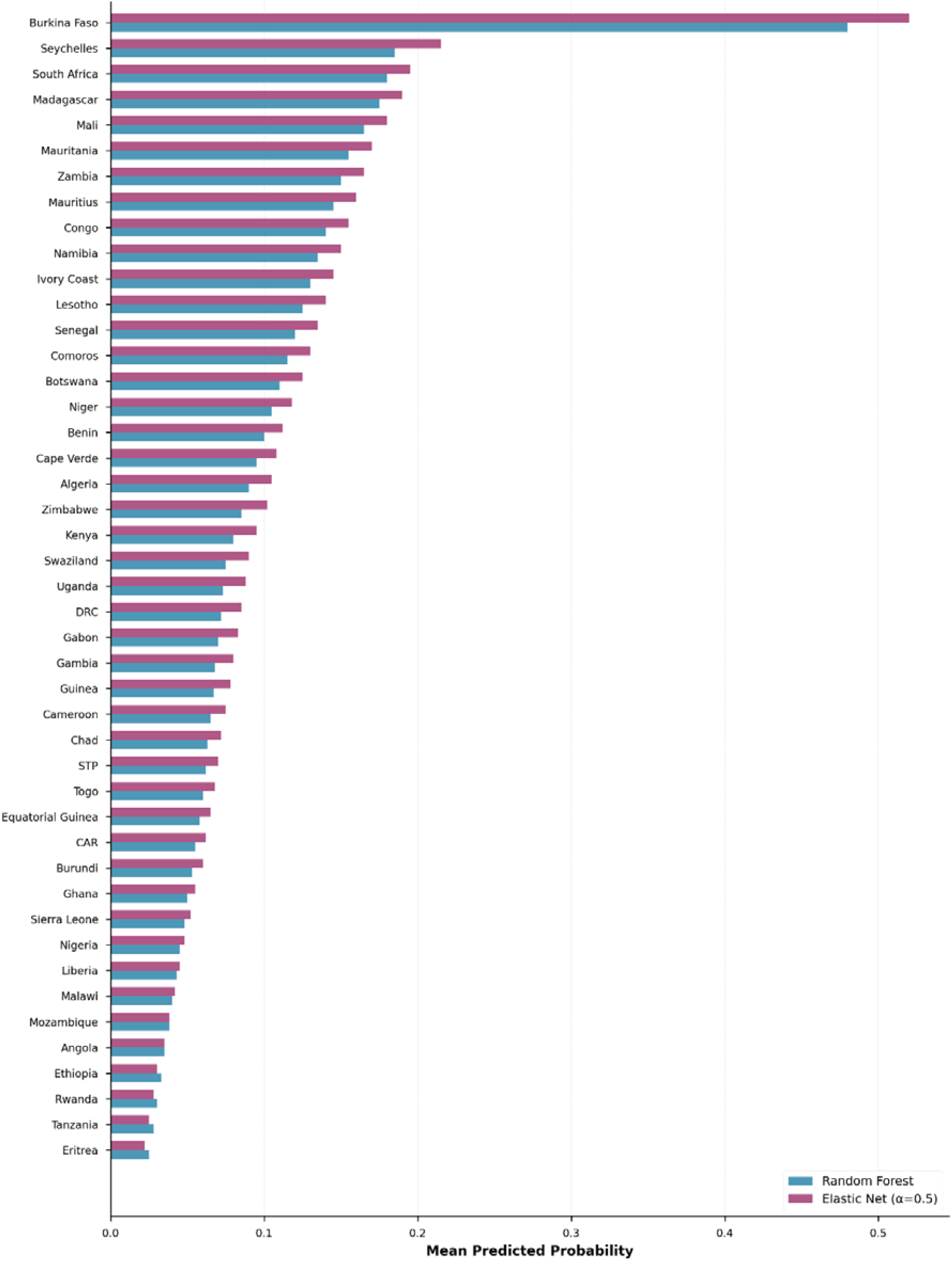
Mean Predicted Smoking Probability in 45 African countries. STP = São Tomé and Príncipe; CAR = Central African Republic

### Smoking probabilities by age group and sex

As shown in figure 2 below, the comparison of the mean predicted probability by gender using the Random Forest and the Elastic Net (α=0.5) predictive models indicated a consistent pattern between the two with males showing higher probability than females. Using the Elastic Net, the predicted mean for males was 0.14 while that of females was 0.07. Similarly, the predicted mean for males was about 0.135 versus 0.065 for females using the Random Forest. The production of almost identical predictions by the models within each sex category, reviewed a strong alignment between the modelling techniques. This implies that sex is an important predictor in the models. The predicted probabilities increased steadily with age reaching a peak between 16 and 18 years (Elastic Net 0.143 vs 0.135 Random Forest), followed by 14-15 and finally the 11-13 years. The increase is noticeable in both models but it is slightly higher in the Elastic Net model in the older age group. The observed increase in prediction values with age indicated that the predicted outcome was highly likely with advancing age. Regionally, Southern Africa demonstrated the highest predicted probability, followed by West Africa, Central Africa, and East Africa. As observed under gender and age demographics, the Elastic Net model produced slightly higher probabilities than the Random Forest across regions.

**Figure 2:**
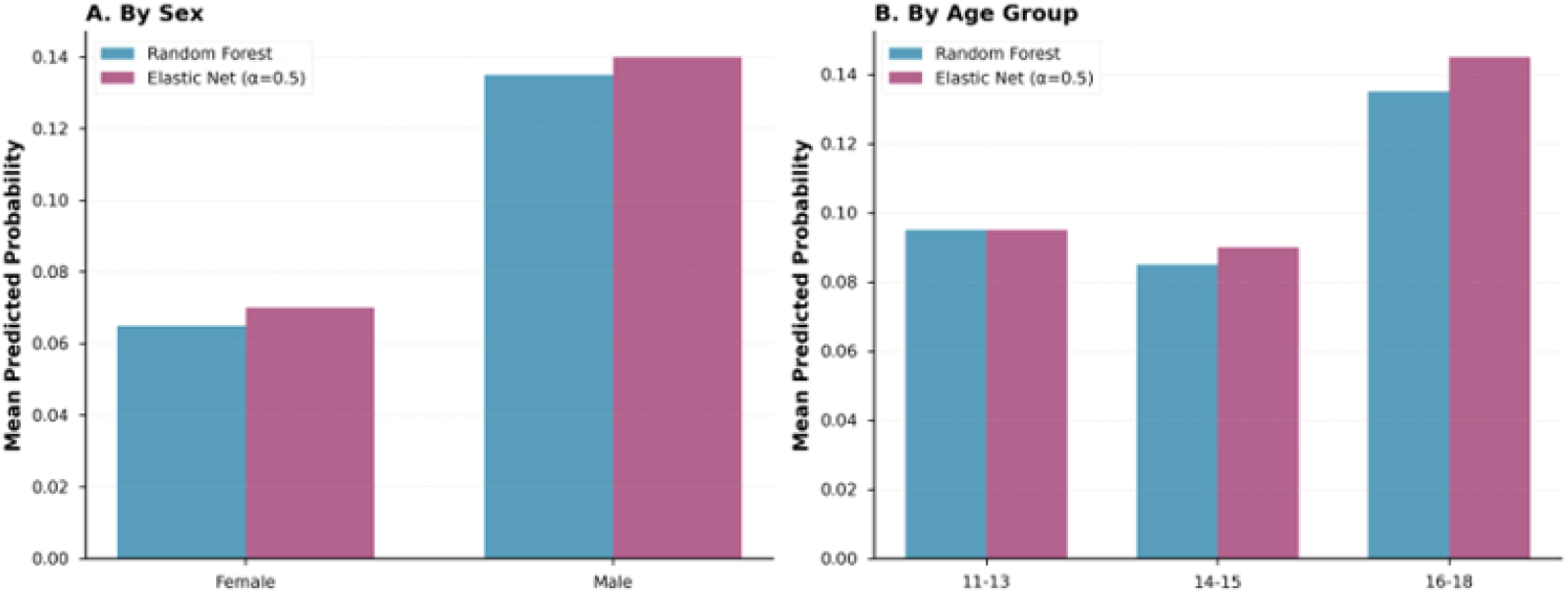
Predictive Model Performance Comparison Across Demographics in 45 African Countries.

Regarding model prediction differences, the distribution of prediction was concentrated around zero (0.00) with the majority of values clustered around the no difference line. This showed a strong consensus between the two predictive models at individual prediction level. The almost symmetrical spread of values around zero for the two techniques, indicated they both yielded comparable predictive outputs without under or over predicting relative to the other.

### Smoking probabilities by age-sex groups

Figure 3 illustrates the predicted probability of cigarette smoking among adolescents aged 11 to 18 years old disaggregated by gender using Random Forest and Elastic Net (α=0.5) predictive models. Among female adolescents, the predicted probability was low in early adolescence and steadily declined between the ages of 13 and 14 before gradually increasing. The sharpest increase in both models was observed between the ages of 16 and 18. The predictive models produced comparable estimates for both males and females across all ages indicating a consistent pattern prediction regardless of the modeling approach used. The similarity between curves among female adolescents indicates that age-related variations in smoking risk is strong and not model-dependent. The predicted probability among males was uniformly higher than that observed among female adolescents. It initially declined in early adolescence (12-13 years) before steadily rising from age 15.

**Figure 3:**
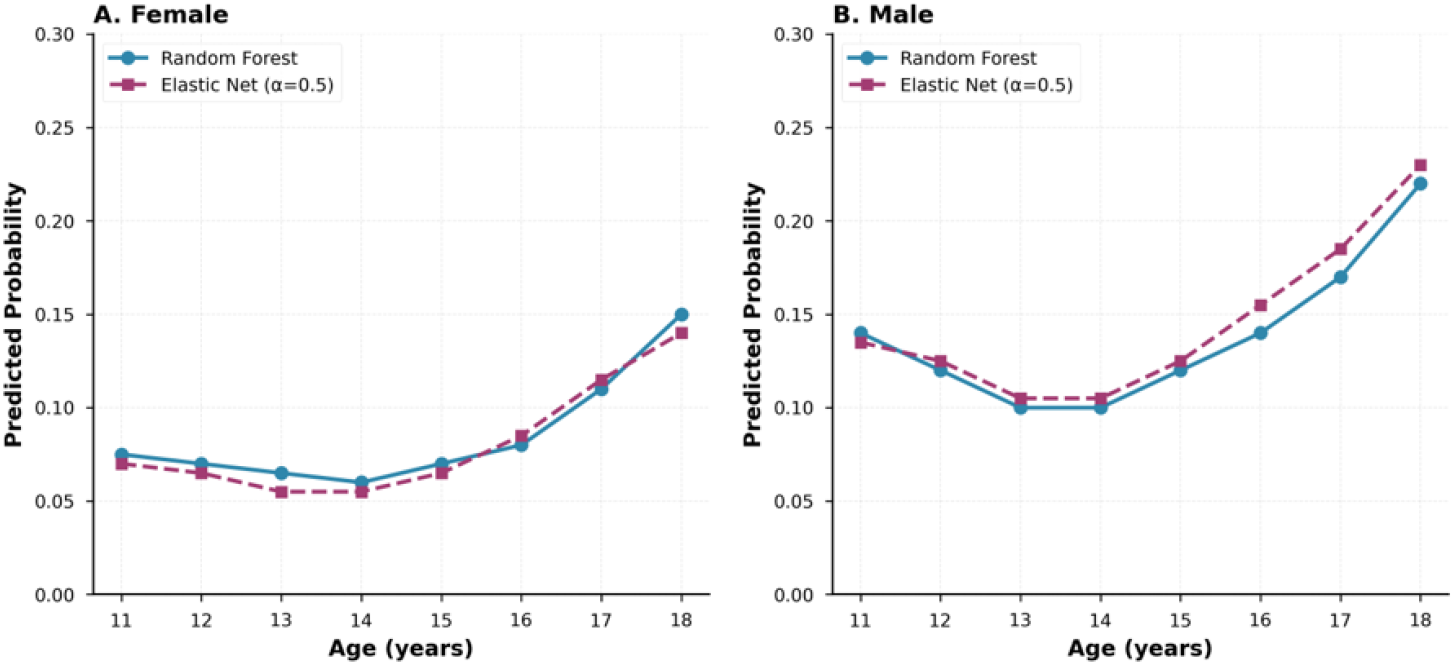
Predicted Smoking Probability by Age and Sex in 45 African countries: 2001-2021.

The sharpest rise was observed between age 17 and 18. Both predictive models showed a uniform course, although the Elastic Net predictions were higher at older ages.

In general, the pattern observed reviewed a robust age-related increase of cigarette smoking risk among males than females, with close alignment of the two modelling techniques.

## DISCUSSION

This study offers important insights into cigarette smoking among school-going adolescents in 45 African countries. The results reviewed that the smoking prevalence and the likelihood of smoking varied widely across countries ranging from 1.5% in Angola to 48.9% in Burkina Faso. We found a pooled prevalence of 23.9%. Because the Global Youth Tobacco Survey focuses only on school going adolescents, this figure may not reflect the true population exposure. The results also showed a rise in cigarette smoking with increasing age peaking at 18 years (22.4%). A similar trend was observed with an increase in school grade. The highest smoking prevalence was observed among students in grade 13 (22.8%). Overall, male (14.1%) participants were much more likely to smoke than their female counterparts (7.3%). The use of other tobacco products such as chew, snuff, or shisha was strongly associated with cigarette smoking. Additionally, ownership of branded tobacco items emerged as a significant factor with participants who owned branded tobacco items more likely to smoke than those who didn’t. Adolescents who were exposed to smoking advertising had a lower prevalence compared to those who were not (10.8% vs 23.8%). Exposure to anti-smoking in the media was associated with a higher smoking prevalence than non-exposure. Students who were taught in school about the effects of tobacco use exhibited a much lower prevalence than those who were not. Notably, students who supported banning of tobacco sales to minors, tobacco advertising and tobacco smoking in public had a lower prevalence of cigarette smoking compared to their counterparts. Awareness of the tobacco control act was associated with lower smoking prevalence among the respondents. These findings offer critical insights into the prevalence and predictors of cigarette smoking among school-going adolescents across 45 countries in Africa.

The finding that 23.9% of students in the 45 countries currently smoked cigarettes falls outside the global estimates from the World Health Organization, which report youth smoking rates ranging between 5.0% and 20.0% in various regions (27). This high prevalence is alarming and inconsistent with most of the studies done globally (23,28). However, a study done in Eastern Ethiopia among adolescents equally had a prevalence rate above 20% (21%) (29). This high cigarette smoking result could be attributed to aggressive and innovative tobacco marketing strategies employed by tobacco companies in Africa that appeal to the youth (30).

The increase in cigarette smoking with age (31) and school grade mirrors patterns seen worldwide (32,33). This rise is influenced by peer pressure, stress related to social transitions and increasing autonomy (34,35). These findings underscore the need to understand factors driving youth tobacco initiation and progression to regular smoking.

The predominance of cigarette smoking among male students observed in the study, reflects a consistent pattern seen both globally and across Africa (14,36,37). Cultural norms and gender roles often influence tobacco use, with males more socially permitted to smoke (38). These findings emphasize the need for gender-sensitive tobacco control policies and interventions that address male social norms and peer influences, in line with global calls to reduce gender disparities in tobacco use (39).

The association between other tobacco product use and cigarette smoking reflects global evidence on dual and poly-tobacco use among youth (40). Adolescents who have used non-cigarette tobacco products were more likely to initiate cigarette smoking (41). A prospective cohort study of the Population Assessment of Tobacco and Health involving over 10,000 youths in the US reviewed that youths who used non-cigarette products were more likely to initiate cigarette smoking a year later as compared to youths who had never used any tobacco products (42). A number of factors could explain this association. Other tobacco products might cause nicotine dependence (43) which some youths perceive might only be satisfied by conventional cigarettes. Additionally, youths might consider cigarettes to be more convenient than other tobacco products (42). This suggests that tobacco control and prevention programs need a comprehensive approach, addressing all nicotine and tobacco products rather than focusing solely on cigarettes (44,45). The study supports the World Health Organization Framework Convention on Tobacco Control’s (WHO FCTC) emphasis on multi-sectoral and integrated tobacco control strategies (46).

The association between ownership of branded tobacco items and smoking was strong in our study. Previous research has shown that ownership of branded tobacco items was associated with cigarette smoking (47,48). Branding of items is part of a strategy utilized by tobacco multinational companies called TAPS (tobacco advertising, promotion and sponsorship) (47). Exposure to TAPS has been shown to increase tobacco initiation and continued use among the youth (47,48). In our study, adolescents who were exposed to cigarette smoking advertising had a lower smoking prevalence while those exposed to anti-smoking advertising in the media surprisingly had a higher smoking prevalence. This finding is unexpected, as it contrasts with the majority of existing literature, which reports higher smoking rates among adolescents exposed to tobacco advertising and lower smoking rates for adolescents exposed to anti-smoking messages (47,48). The observed trend may be attributable to random variation, but it also highlights the need for further investigation into the underlying factors driving this pattern.

Being taught in school about the effects of tobacco use emerged as protective factor against cigarette smoking, with students who were taught about smoking reporting lower smoking prevalence. This is consistent with other studies done in Africa (17).

In our study, students who supported banning of tobacco sales to minors, tobacco advertising and smoking in public had lower smoking prevalence which conforms with findings from other studies (17,35,38).

Despite advertising bans in many countries, indirect marketing continues to pose a challenge (49,50). The findings highlight the urgent need to close regulatory loopholes in tobacco advertising, promotion, and sponsorship laws in Africa, supporting global efforts under the Framework Convention on Tobacco Control (FCTC) to protect youth from tobacco marketing.

### Strengths and limitations

There are limitations to the study that should be considered when interpreting the results. First, this study relies on self-reported data, which has the potential to introduce reporting bias, particularly under-reporting of smoking due to social desirability resulting in possible underestimation of the actual prevalence. Secondly, as this was an observational study, we were able to identify factors linked to cigarette smoking among school-going adolescents; however, causal relationships could not be established. While the multivariable model controlled for recognized confounders, residual confounding, often present in observational studies, may still exist and could influence the interpretation of the findings. Thirdly, the cross-sectional nature of the study, limits generalizability of the findings to other international contexts. Additionally, variability in sample sizes and data collection years across countries may affect comparability.

Despite the limitations, the study offers some strengths. The study is drawn from a large continent-wide dataset spanning two decades with more than 400,000 school going adolescents across 45 African countries. It therefore provides a comprehensive overview of the prevalence and predictors of cigarette smoking among school-going adolescents addressing a significant gap in public health literature. By integrating traditional and predictive approaches in the analysis, our study was able to describe and explain observed associations and predict the probability of cigarette smoking among school-going adolescents in the future, thereby informing progressive decision-making. The study offers important insights to guide public health intervention and policy development. In addition, the application of rigorous statistical methods, with adjustments for confounders, strengthens the credibility and robustness of the results.

## CONCLUSION

In conclusion, this study offers valuable large-scale, multi-country evidence on cigarette smoking among students in Africa, revealing key determinants and substantial variations across different contexts. The findings underscore the urgent need for targeted, context-specific prevention efforts - particularly those focusing on early adolescence, male youth, and the implementation of comprehensive tobacco control policies that address all tobacco products and marketing. Strengthening tobacco control among students is essential to reduce the long-term health burden of tobacco use and to advance progress toward the Sustainable Development Goals related to health and non-communicable diseases.

## Supporting information

Supplemental codes

## ABBREVIATIONS

AIC: Akaike Information Criterion
AUC: Area Under the Receiver Operating Characteristic Curve
CDC: Center for Disease Control and Prevention
CI: Confidence Interval
FCTC: Framework Convention on Tobacco Control
GYTS: Global Youth Tobacco Survey
IVF: Variance Inflation Factor
TAPS: Tobacco Advertising Promotion and Sponsorship
US: United States
WHO: World Health Organization

## Author’s contributions

JS performed data management, analysis and drafting the paper; WN and CZ reviewed the data management, analysis and the draft paper. All authors approved the final draft manuscript.

## Data availability

The datasets analysed during the current study were obtained from the Global Youth Tobacco Surveys and are publicly available. Access to the raw data can be requested from the World Health Organization (WHO) through the WHO NCD Microdata Repository or by contacting the WHO NCD Surveillance Team at, subject to approval and relevant data-sharing agreements. The analytical code used in this study is available from the author upon reasonable request.

## Declarations

### Ethics approval and consent to participate

Ethical approval was obtained from the University of Zambia Biomedical Research Ethics Committee (Ref. No.7265-2025) and the National Health Research Authority of Zambia (Ref. No: NHRA-2899/14/11/2025), in compliance with national research guidelines. Access to the anonymized GYTS datasets was granted by the WHO through the NCD Microdata Repository. As the study involved no direct contact with participants and used de identified data, no additional consent was required.

This study is a secondary analysis of de-identified GYTS data. The authors accessed the data through an approved data request to the World Health Organization. The analysis involved no direct contact with participants and did not require additional ethical approval, as the datasets were fully anonymized.

### Consent for publication

This manuscript does not contain any individual-level identifiable data and therefore Consent for publication is not applicable.

### Funding information

We did not receive any funding to conduct this analysis.

### Competing interest

We declare that there was no competing interest. The paper was prepared as part of the Doctoral Studies in Global Health at University of Zambia.

## Notes

### Competing Interest Statement

The authors have declared no competing interest.

### Funding Statement

None

## REFERENCES

1. World Health Organization. WHO report on the global tobacco epidemic, 2025: warning about the dangers of tobacco.

2. Belo R Sofia, Elif D, Paraskevi K, E L Keir, Charlotta P. Supporting Tobacco Cessation. European Respiratory Society; 2021. 327 p.

3. Sakthisankaran SM, Sakthipriya D, Swamivelmanickam M. Health Risks Associated with Tobacco Consumption in Humans: An Overview. J Drug Deliv Ther [Internet]. 2024 May 15 [cited 2025 Jul 7];14(5):163–73. Available from: https://jddtonline.info/index.php/jddt/article/view/6523

4. Chugh A, Jain N, Arora M. Prevention and Control of Tobacco Use as a Major Risk Factor for Non-communicable Diseases (NCDS): A Lifecourse Approach. In: Kelishadi R, editor. Healthy Lifestyle [Internet]. Cham: Springer International Publishing; 2022 [cited 2025 Jul 7]. p. 173–97. (Integrated Science; vol. 3). Available from: https://link.springer.com/10.1007/978-3-030-85357-0_9

5. World Health Organization. Tobacco and its environmental impact: an overview [Internet]. Geneva: World Health Organization; 2017 [cited 2025 Jul 7]. 72 p. Available from: https://iris.who.int/handle/10665/255574

6. Carreras G, Lugo A, Gallus S, Cortini B, Fernández E, López MJ, et al. Burden of disease attributable to second-hand smoke exposure: A systematic review. Prev Med [Internet]. 2019 Dec [cited 2025 Jul 7];129:105833. Available from: https://linkinghub.elsevier.com/retrieve/pii/S0091743519303093

7. Monyeki KD, Kemper HCG. Lifestyle and Epidemiology: The Double Burden of Poverty and Cardiovascular Diseases in African Populations. BoD – Books on Demand; 2021. 336 p.

8. Chileshe B, Ng’ambi WF, Habbanti SG, Zyambo C. Policy implication of high prevalence of tobacco use and alcohol consumption among school-going adolescents in 2021: A cross-sectional study in a mining town of Kalulushi Zambia. Int J Noncommunicable Dis [Internet]. 2025 Jan [cited 2025 Dec 18];10(1):17–24. Available from: https://journals.lww.com/10.4103/jncd.jncd_123_24

9. Casetta B, Videla AJ, Bardach A, Morello P, Soto N, Lee K, et al. Association Between Cigarette Smoking Prevalence and Income Level: A Systematic Review and Meta-Analysis. Nicotine Tob Res [Internet]. 2016 Sep 27 [cited 2025 Jul 7];ntw266. Available from: https://academic.oup.com/ntr/article-lookup/doi/10.1093/ntr/ntw266

10. Ogbodo SC, Onyekwum CA. Social determinants of health, religiosity, and tobacco use in sub-Saharan Africa: evidence from the global adult tobacco surveys in seven countries. J Public Health [Internet]. 2024 Jun [cited 2025 Jul 7];32(6):895–908. Available from: https://link.springer.com/10.1007/s10389-023-01882-9

11. Immurana M, Boachie MK, Iddrisu AA. The effects of tobacco taxation and pricing on the prevalence of smoking in Africa. Glob Health Res Policy [Internet]. 2021 Dec [cited 2025 Jul 7];6(1):14. Available from: https://ghrp.biomedcentral.com/articles/10.1186/s41256-021-00197-0

12. Reitsma MB, Fullman N, Ng M, Salama JS, Abajobir A, Abate KH, et al. Smoking prevalence and attributable disease burden in 195 countries and territories, 1990–2015: a systematic analysis from the Global Burden of Disease Study 2015. The Lancet [Internet]. 2017 May [cited 2025 Jul 7];389(10082):1885–906. Available from: https://linkinghub.elsevier.com/retrieve/pii/S014067361730819X

13. Oyewole BK, Animasahun VJ, Chapman HJ. Tobacco use in Nigerian youth: A systematic review. Wang W, editor. PLOS ONE [Internet]. 2018 May 3 [cited 2025 Jul 8];13(5):e0196362. Available from: https://dx.plos.org/10.1371/journal.pone.0196362

14. James PB, Bah AJ, Kabba JA, Kassim SA, Dalinjong PA. Prevalence and correlates of current tobacco use and non-user susceptibility to using tobacco products among school-going adolescents in 22 African countries: a secondary analysis of the 2013-2018 global youth tobacco surveys. Arch Public Health [Internet]. 2022 Dec [cited 2025 Jul 7];80(1):121. Available from: https://archpublichealth.biomedcentral.com/articles/10.1186/s13690-022-00881-8

15. World Health Organization. WHO NCD microdata repository [Internet]. 2023 [cited 2025 Jul 21]. Available from: https://extranet.who.int/ncdsmicrodata/index.php/catalog/?page=1&ps=15

16. Aboagye RG, Mohammed A, Duodu PA, Adnani QES, Seidu AA, Ahinkorah BO. Sex-related inequalities in current cigarette smoking among adolescents in Africa. Subst Abuse Treat Prev Policy [Internet]. 2024 Sep 5 [cited 2025 Jul 7];19(1):41. Available from: https://substanceabusepolicy.biomedcentral.com/articles/10.1186/s13011-024-00619-5

17. Zyambo C, Olowski P, Mulenga D, Liamba F, Syapiila P, Siziya S. School tobacco-related curriculum and behavioral factorsassociated with cigarette smoking among school-goingadolescents in Zambia: Results from the 2011 GYTS study. Tob Induc Dis [Internet]. 2022 May 5 [cited 2025 Jul 9];20(May):1–9. Available from: http://www.tobaccoinduceddiseases.org/School-tobacco-related-curriculum-and-behavioral-factors-nassociated-with-cigarette,146960,0,2.html

18. Papaleontiou L, Agaku IT, Filippidis FT. Effects of Exposure to Tobacco and Electronic Cigarette Advertisements on Tobacco Use: An Analysis of the 2015 National Youth Tobacco Survey. J Adolesc Health [Internet]. 2020 Jan [cited 2025 Jul 7];66(1):64–71. Available from: https://linkinghub.elsevier.com/retrieve/pii/S1054139X1930312X

19. Peer N. Current strategies are inadequate to curb the rise of tobacco use in Africa. S Afr Med J [Internet]. 2018 Aug 5 [cited 2025 Jul 7];108(7). Available from: https://www.ajol.info/index.php/samj/article/view/175692

20. Egbe CO, Magati P, Wanyonyi E, Sessou L, Owusu-Dabo E, Ayo-Yusuf OA. Landscape of tobacco control in sub-Saharan Africa. Tob Control [Internet]. 2022 Mar [cited 2025 Jul 7];31(2):153–9. Available from: https://tobaccocontrol.bmj.com/lookup/doi/10.1136/tobaccocontrol-2021-056540

21. Sargent JD, Gabrielli J, Budney A, Soneji S, Wills TA. Adolescent smoking experimentation as a predictor of daily cigarette smoking. Drug Alcohol Depend [Internet]. 2017 Jun [cited 2025 Oct 10];175:55–9. Available from: https://linkinghub.elsevier.com/retrieve/pii/S0376871617301370

22. Wellman RJ, Dugas EN, Dutczak H, O’Loughlin EK, Datta GD, Lauzon B, et al. Predictors of the Onset of Cigarette Smoking. Am J Prev Med [Internet]. 2016 Nov [cited 2025 Oct 10];51(5):767–78. Available from: https://linkinghub.elsevier.com/retrieve/pii/S0749379716300940

23. Reitsma MB, Flor LS, Mullany EC, Gupta V, Hay SI, Gakidou E. Spatial, temporal, and demographic patterns in prevalence of smoking tobacco use and initiation among young people in 204 countries and territories, 1990–2019. Lancet Public Health [Internet]. 2021 Jul 1 [cited 2025 Oct 7];6(7):e472–81. Available from: https://www.thelancet.com/journals/lanpub/article/PIIS2468-2667(21)00102-X/abstract

24. Global Youth Tobacco Survey Collaborative Group. Global Youth Tobacco Survey (GYTS): Sample Design and Weights. [Internet]. Atlanta, GA: Centers for Disease Control and Prevention; 2014 [cited 2025 Oct 10]. Report No.: Version 1.1. Available from: https://drupal.gtssacademy.org/wp-content/uploads/2025/06/4GYTS-Sample-Design-and-Weights-Manual.pdf

25. Ng’ambi WF, Muula AS. A reproducible R workflow to preserve variable and value labels in Stata, SPSS, and SAS datasets for transparent and reproducible health research. Malawi Med J [Internet]. 2025 Sep 15 [cited 2025 Dec 28];37(3):193. Available from: https://pmc.ncbi.nlm.nih.gov/articles/PMC12547319/

26. Ng’ambi WF, Zyambo C, Muula AS. A Practical Guide to Key Considerations in Logistic Regression for Clinical and Public Health Research: R tutorial. [cited 2025 Dec 28]; Available from: https://www.mmj.mw/?p=13527

27. World Health Organization. WHO global report on trends in prevalence of tobacco use 2000–2030 [Internet]. Geneva; 2024 [cited 2025 Aug 8]. Available from: https://books.google.com/books/about/WHO_global_report_on_trends_in_prevalenc.html?id=k6YOEQAAQBAJ

28. Itanyi IU, Onwasigwe CN, McIntosh S, Bruno T, Ossip D, Nwobi EA, et al. Disparities in tobacco use by adolescents in southeast, Nigeria using Global Youth Tobacco Survey (GYTS) approach. BMC Public Health [Internet]. 2018 Dec [cited 2025 Jul 8];18(1):1–11. Available from: https://link.springer.com/article/10.1186/s12889-018-5231-1

29. Roble AK, Osman MO, Lathwal OP, Aden AA. Prevalence of Cigarette Smoking and Associated Factors Among Adolescents in Eastern Ethiopia, 2020. Subst Abuse Rehabil [Internet]. 2021 Oct [cited 2026 Feb 18];Volume 12:73–80. Available from: https://www.dovepress.com/prevalence-of-cigarette-smoking-and-associated-factors-among-adolescen-peer-reviewed-fulltext-article-SAR

30. Krishnan-Sarin S, O’Malley SS, Green BG, Jordt SE. The science of flavour in tobacco products. World Health Organ Tech Rep Ser [Internet]. 2019 Oct 24 [cited 2025 Aug 30];1015:125. Available from: https://pmc.ncbi.nlm.nih.gov/articles/PMC9896977/

31. Zyambo C, Olowski P, Mulenga D, Syapiila P, Liwewe MM, Hazemba A, et al. Prevalence and Factors Associated with Tobacco Smoking in a National Representative Sample of Zambian Adults: Data from the 2017 STEPS – NCDs Survey. Asian Pac J Cancer Prev APJCP [Internet]. 2023 [cited 2025 Dec 28];24(1):111. Available from: https://pmc.ncbi.nlm.nih.gov/articles/PMC10152846/

32. Kasza KA, Ambrose BK, Conway KP, Borek N, Taylor K, Goniewicz ML, et al. The New England Journal of Medicine. Massachusetts Medical Society; 2017 [cited 2025 Oct 7]. Tobacco-Product Use by Adults and Youths in the United States in 2013 and 2014. Available from: https://www.nejm.org/doi/full/10.1056/NEJMsa1607538

33. Wellman RJ, Dugas EN, Dutczak H, O’Loughlin EK, Datta GD, Lauzon B, et al. Predictors of the Onset of Cigarette Smoking. Am J Prev Med [Internet]. 2016 Nov [cited 2025 Oct 7];51(5):767–78. Available from: https://linkinghub.elsevier.com/retrieve/pii/S0749379716300940

34. Defoe IN, Semon Dubas J, Somerville LH, Lugtig P, Van Aken MAG. The unique roles of intrapersonal and social factors in adolescent smoking development. Dev Psychol [Internet]. 2016 Dec [cited 2025 Oct 7];52(12):2044–56. Available from: https://doi.apa.org/doi/10.1037/dev0000198

35. Syapiila P, Mulenga D, Mazaba M, Njunju E, Zyambo C, Chongwe G, et al. Factors associated with intention to smoke cigarettes among never smoker school going adolescents in Zambia. Afr Health Sci [Internet]. 2023 Apr 11 [cited 2025 Dec 28];23(1):596–605. Available from: https://www.ajol.info/index.php/ahs/article/view/245612

36. Agaku IT, Sulentic R, Dragicevic A, Njie G, Jones CK, Odani S, et al. Gender differences in use of cigarette and non-cigarette tobacco products among adolescents aged 13–15 years in 20 African countries. Tob Induc Dis [Internet]. 2024 [cited 2025 Jul 8];22:10–18332. Available from: https://pmc.ncbi.nlm.nih.gov/articles/PMC10801700/

37. Luputa C, Ng’ambi WF, Zyambo C. Cigarette availability and affordability among school-going adolescent smokers in Zambia: Results from the 2011 Global Youth Tobacco Survey. Int J Noncommunicable Dis [Internet]. 2024 Jul [cited 2025 Dec 28];9(3):126–33. Available from: https://journals.lww.com/10.4103/jncd.jncd_49_24

38. Pokothoane R, Agerfa TG, Miderho CC, Mdege ND. Prevalence and determinants of tobacco use among school-going adolescents in 53 African countries: Evidence from the Global Youth Tobacco Survey. Addict Behav Rep [Internet]. 2025 Jun 1 [cited 2025 Jul 7];21:100581. Available from: https://www.sciencedirect.com/science/article/pii/S2352853224000580

39. Lahiri S, Bingenheimer JB, Evans WD, Wang Y, Dubey P, Snowden B. Social Norms Change and Tobacco Use: A Protocol for a Systematic Review and Meta-Analysis of Interventions. Int J Environ Res Public Health [Internet]. 2021 Nov 20 [cited 2025 Aug 5];18(22):12186. Available from: https://www.mdpi.com/1660-4601/18/22/12186

40. Chen DTH, Girvalaki C, Mechili EA, Millett C, Filippidis FT. Global Patterns and Prevalence of Dual and Poly-Tobacco Use: A Systematic Review. [cited 2025 Jul 8]; Available from: 10.1093/ntr/ntab084

41. Stokes AC, Wilson AE, Lundberg DJ, Xie W, Berry KM, Fetterman JL, et al. Racial/Ethnic Differences in Associations of Non-cigarette Tobacco Product Use With Subsequent Initiation of Cigarettes in US Youths. Nicotine Tob Res [Internet]. 2021 May 24 [cited 2025 Oct 6];23(6):900–8. Available from: https://academic.oup.com/ntr/article/23/6/900/5908776

42. Watkins SL, Glantz SA, Chaffee BW. Association of Noncigarette Tobacco Product Use With Future Cigarette Smoking Among Youth in the Population Assessment of Tobacco and Health (PATH) Study, 2013-2015. JAMA Pediatr [Internet]. 2018 Feb 1 [cited 2025 Oct 6];172(2):181. Available from: http://archpedi.jamanetwork.com/article.aspx?doi=10.1001/jamapediatrics.2017.4173

43. Bahelah R, DiFranza JR, Fouad FM, Ward KD, Eissenberg T, Maziak W. Early symptoms of nicotine dependence among adolescent waterpipe smokers. Tob Control [Internet]. 2016 Dec [cited 2025 Oct 6];25(e2):e127–34. Available from: https://tobaccocontrol.bmj.com/lookup/doi/10.1136/tobaccocontrol-2015-052809

44. Bhatnagar A, Whitsel LP, Blaha MJ, Huffman MD, Krishan-Sarin S, Maa J, et al. New and Emerging Tobacco Products and the Nicotine Endgame: The Role of Robust Regulation and Comprehensive Tobacco Control and Prevention: A Presidential Advisory From the American Heart Association. Circulation [Internet]. 2019 May 7 [cited 2025 Jul 8];139(19). Available from: https://www.ahajournals.org/doi/10.1161/CIR.0000000000000669

45. Sanni S, Hongoro C, Ndinda C, Wisdom JP. Assessment of the multi-sectoral approach to tobacco control policies in South Africa and Togo. BMC Public Health [Internet]. 2018 Aug [cited 2025 Jul 8];18(1):1–12. Available from: https://link.springer.com/article/10.1186/s12889-018-5829-3

46. Lencucha R, Reddy SK, Labonte R, Drope J, Magati P, Goma F, et al. Global tobacco control and economic norms: an analysis of normative commitments in Kenya, Malawi and Zambia. Health Policy Plan [Internet]. 2018 Apr 1 [cited 2025 Dec 28];33(3):420–8. Available from: https://academic.oup.com/heapol/article/33/3/420/4833987

47. Chido-Amajuoyi OG, Mantey DS, Clendennen SL, Pérez A. Association of tobacco advertising, promotion and sponsorship (TAPS) exposure and cigarette use among Nigerian adolescents: implications for current practices, products and policies. BMJ Glob Health [Internet]. 2017 Aug [cited 2025 Aug 30];2(3):e000357. Available from: https://gh.bmj.com/lookup/doi/10.1136/bmjgh-2017-000357

48. Vallone D, Greenberg M, Xiao H, Bennett M, Cantrell J, Rath J, et al. The Effect of Branding to Promote Healthy Behavior: Reducing Tobacco Use among Youth and Young Adults. Int J Environ Res Public Health [Internet]. 2017 Dec [cited 2025 Jul 8];14(12):1517. Available from: https://www.mdpi.com/1660-4601/14/12/1517

49. Husain MJ, English LM, Ramanandraibe N. An overview of tobacco control and prevention policy status in Africa. Prev Med [Internet]. 2016 Oct [cited 2025 Jul 8];91:S16–22. Available from: https://linkinghub.elsevier.com/retrieve/pii/S0091743516000645

50. Egbe CO, Bialous SA, Glantz SA. Avoiding “A Massive Spin-Off Effect in West Africa and Beyond”: The Tobacco Industry Stymies Tobacco Control in Nigeria. Nicotine Tob Res [Internet]. 2017 Jul 1 [cited 2025 Jul 8];19(7):877–87. Available from: https://academic.oup.com/ntr/article/19/7/877/2990236

