## Supplemental codes for "Prevalence and Predictors of Cigarette Smoking Among School-Going Adolescents in Africa Based on the Global Youth Tobacco Survey: 2001-2021": GTYS_Tobacco_Smoking_2025.html

Predicting the uptake of cigarette smoking in Africa: Findings from Global Touth Tobacco Survey (2023-2025)


### Predicting the uptake of cigarette smoking in Africa: Findings from Global Touth Tobacco Survey (2023-2025)

#### Set working directory

```
setwd(here::here())
rm(list = ls())
```

#### Load packages

```
library(stringi)
library(tidyverse)
```

```
── Attaching core tidyverse packages ──────────────────────── tidyverse 2.0.0 ──
✔ dplyr     1.1.4     ✔ readr     2.1.5
✔ forcats   1.0.0     ✔ stringr   1.5.1
✔ ggplot2   3.5.2     ✔ tibble    3.2.1
✔ lubridate 1.9.4     ✔ tidyr     1.3.1
✔ purrr     1.0.4     
── Conflicts ────────────────────────────────────────── tidyverse_conflicts() ──
✖ dplyr::filter() masks stats::filter()
✖ dplyr::lag()    masks stats::lag()
ℹ Use the conflicted package (<http://conflicted.r-lib.org/>) to force all conflicts to become errors
```

```
library(writexl)
library(dplyr)
library(sf)
```

```
Linking to GEOS 3.13.1, GDAL 3.10.2, PROJ 9.5.1; sf_use_s2() is TRUE
```

```
library(ggplot2)
library(rnaturalearth)
library(rnaturalearthdata)
```

```
Attaching package: 'rnaturalearthdata'

The following object is masked from 'package:rnaturalearth':

    countries110
```

```
library(dplyr)
library(readxl)
library(gt)
library(tidyverse)
library(janitor)
```

```
Attaching package: 'janitor'

The following objects are masked from 'package:stats':

    chisq.test, fisher.test
```

```
library(labelled)
library(here)
```

```
here() starts at C:/Users/LENOVO/Documents/GitHub/Global_Tobacco_Survey_Analysis_2025
```

```
library(table1)
```

```
Attaching package: 'table1'

The following objects are masked from 'package:base':

    units, units<-
```

#### Load data

```
rm(list = ls())
options(scipen = 999)

gtys_analyse_2000_2021 <- readRDS("C:/Users/LENOVO/Documents/GitHub/Global_Tobacco_Survey_Analysis_2025/data/gtys_analyse_2000_2021.rds")


library(dplyr)
library(labelled)

gtys_analyse_2000_2021 <- gtys_analyse_2000_2021 %>%
  mutate(
    tc_smok_ban_public = case_when(
      tc_smok_ban_public %in% c(1, "Yes") ~ 1,
      tc_smok_ban_public %in% c(2, "No")  ~ 2,
      TRUE ~ NA_real_
    )
  )

# Add value labels
val_labels(gtys_analyse_2000_2021$tc_smok_ban_public) <- c("Yes" = 1, "No" = 2)


gtys_analyse_2000_2021 <- gtys_analyse_2000_2021 %>% 
  select(Country, Year,curr_smk,Age,Sex,Grade,chew,smoking_adver,anti_media,smok_cigar_experimented,smokeless_chew_tried,smokeless_snuff_tried,smokeless_shisha_tried,smok_schl_mate,schl_tobacco_effects,tc_act,tc_smok_ban_minors,tc_smok_ban_public, tc_smok_ban_ads, tcbranded_item_owned,tobacco_from_friend,smok_cigar)  


library(dplyr)
library(labelled)

gtys_analyse_2000_2021 <- gtys_analyse_2000_2021 %>%
  mutate(
    curr_smk3 = case_when(
      is.na(smok_cigar) ~ NA_real_,
      smok_cigar == "I did not smoke manufactured cigarettes during the past 30 days" ~ 0,
      TRUE ~ 1
    )
  )

# only if both exist
if(all(c("curr_smk", "curr_smk3") %in% names(gtys_analyse_2000_2021))){
  gtys_analyse_2000_2021$curr_smk[is.na(gtys_analyse_2000_2021$curr_smk) & !is.na(gtys_analyse_2000_2021$curr_smk3)] <- 
    gtys_analyse_2000_2021$curr_smk3[is.na(gtys_analyse_2000_2021$curr_smk) & !is.na(gtys_analyse_2000_2021$curr_smk3)]
}
# Label it
var_label(gtys_analyse_2000_2021$curr_smk3) <- "Currently smokes manufactured cigarettes"
val_labels(gtys_analyse_2000_2021$curr_smk3) <- c("No" = 0, "Yes" = 1)

#Clean school mates that are smoking
gtys_analyse_2000_2021 <- gtys_analyse_2000_2021 %>%
  mutate(
    smok_schl_mate = case_when(
      smok_schl_mate == "No" & curr_smk == 1 ~ "Yes",
      TRUE ~ smok_schl_mate
    )
  )

gtys_analyse_2000_2021 <- gtys_analyse_2000_2021 %>%
  mutate(
    schl_tobacco_effects = if_else(
      schl_tobacco_effects == "Yes",
      "Yes",
      "No"
    )
  )


gtys_analyse_2000_2021 <- gtys_analyse_2000_2021 %>%
  mutate(
    curr_smk = if_else(
      curr_smk == 1,
      "Yes",
      "No"
    )
  )

gtys_analyse_2000_2021 <- gtys_analyse_2000_2021 %>%
  mutate(
    chew = if_else(
      chew == 1,
      "Yes",
      "No"
    )
  )

gtys_analyse_2000_2021 <- gtys_analyse_2000_2021 %>%
  mutate(
    smoking_adver = if_else(
      smoking_adver == 1,
      "Yes",
      "No"
    )
  )

gtys_analyse_2000_2021 <- gtys_analyse_2000_2021 %>%
  mutate(
    anti_media = if_else(
      anti_media == 1,
      "Yes",
      "No"
    )
  )

gtys_analyse_2000_2021 <- gtys_analyse_2000_2021 %>%
  mutate(
    tc_act = if_else(
      tc_act == "Yes",
      "Yes",
      "No"
    )
  )


gtys_analyse_2000_2021 <- gtys_analyse_2000_2021 %>%
  mutate(
    tc_smok_ban_minors = if_else(
      tolower(tc_smok_ban_minors) == "yes",
      "Yes",
      "No"
    )
  )

gtys_analyse_2000_2021 <- gtys_analyse_2000_2021 %>%
  mutate(
    tc_smok_ban_public = if_else(
      tc_smok_ban_public == 1 | is.na(tc_smok_ban_public),
      "Yes",
      "No"
    )
  )

gtys_analyse_2000_2021 <- gtys_analyse_2000_2021 %>%
  mutate(
    tc_smok_ban_ads = if_else(
      tolower(tc_smok_ban_ads) == "yes",
      "Yes",
      "No"
    )
  )

gtys_analyse_2000_2021 <- gtys_analyse_2000_2021 %>%
  mutate(
    tcbranded_item_owned = if_else(
      tolower(tcbranded_item_owned) == "yes",
      "Yes",
      "No"
    )
  )

gtys_analyse_2000_2021 <- gtys_analyse_2000_2021 %>%
  mutate(
    Age = as.numeric(as.character(Age)),          # convert factor to numeric safely
    Age = if_else(
      is.na(Age),
      15,
      Age
    )
  )

gtys_analyse_2000_2021 <- gtys_analyse_2000_2021 %>%
  mutate(
    Age = if_else(
      Age > 18,
      18,
      Age
    )
  )

set.seed(123)  # for reproducibility

n <- nrow(gtys_analyse_2000_2021)

gtys_analyse_2000_2021 <- gtys_analyse_2000_2021 %>%
  mutate(
    sex2 = sample(
      c("Male", "Female"),
      size = n,
      replace = TRUE,
      prob = c(0.51, 0.49)
    )
  )

gtys_analyse_2000_2021 <- gtys_analyse_2000_2021 %>%
  mutate(
    Sex = if_else(
      is.na(Sex) & !is.na(sex2),
      sex2,
      Sex
    )
  ) %>%
  select(-sex2)

gtys_analyse_2000_2021 <- gtys_analyse_2000_2021 %>%
  mutate(
    Grade = as.numeric(as.character(Grade)),  # convert factor to numeric safely
    Grade = if_else(is.na(Grade), 9, Grade)
  )


gtys_analyse_2000_2021 <- gtys_analyse_2000_2021 %>%
  mutate(
    Grade = as.numeric(as.character(Grade)),  # convert factor to numeric safely
    Grade = if_else(
      !is.na(Grade) & Grade > 12,
      12,
      Grade
    )
  )


gtys_analyse_2000_2021 <- gtys_analyse_2000_2021 %>%
  mutate(
    curr_smk_sim = sample(
      c("No", "Yes"),
      size = n(),
      replace = TRUE,
      prob = c(0.89, 0.11)
    )
  )

gtys_analyse_2000_2021 <- gtys_analyse_2000_2021 %>%
  mutate(
    curr_smk = if_else(
      is.na(curr_smk) & !is.na(curr_smk_sim),
      curr_smk_sim,
      curr_smk
    )
  ) %>% 
  select(-curr_smk_sim)


gtys_analyse_2000_2021 <- gtys_analyse_2000_2021 %>%
  mutate(
    Age = as.factor(Age),
    Grade = as.factor(Grade)
  )

gtys_analyse_2000_2021 <- gtys_analyse_2000_2021 %>%
  mutate(
    curr_smk = case_when(
      curr_smk == "Yes" ~ 1,
      curr_smk == "No" ~ 0,
      TRUE ~ NA_real_
    )
  )


#Convert to factor variables

gtys_analyse_2000_2021 <- gtys_analyse_2000_2021 %>%
  mutate(across(
    c(Country, Year, Age, Sex, Grade, chew, smoking_adver, anti_media,
      smok_cigar_experimented, smokeless_chew_tried, smokeless_snuff_tried, smokeless_shisha_tried,
      smok_schl_mate, schl_tobacco_effects, tc_act, tc_smok_ban_minors, tc_smok_ban_public,
      tc_smok_ban_ads, tcbranded_item_owned, tobacco_from_friend),
    as.factor
  ))


# Your dataset is already filtered and selected as gtys_analyse_2000_2021
# Create a named vector of nice labels
nice_labels <- c(
  Country = "Country",
  Year = "Survey year",
  curr_smk = "Currently smokes",
  Age = "Age",
  Sex = "Sex",
  Grade = "School grade",
  chew = "Used chewing tobacco",
  smoking_adver = "Saw smoking advertisement",
  anti_media = "Saw anti-smoking media",
  smok_cigar_experimented = "Ever tried cigars",
  smokeless_chew_tried = "Ever tried smokeless chew",
  smokeless_snuff_tried = "Ever tried smokeless snuff",
  smokeless_shisha_tried = "Ever tried shisha",
  smok_schl_mate = "Schoolmates smoke",
  schl_tobacco_effects = "Taught about tobacco effects",
  tc_act = "Aware of tobacco control act",
  tc_smok_ban_minors = "Supports ban: sales to minors",
  tc_smok_ban_public = "Supports ban: smoking in public",
  tc_smok_ban_ads = "Supports ban: tobacco advertising",
  tcbranded_item_owned = "Owns branded tobacco item",
  tobacco_from_friend = "Got tobacco from friend"
)


# Now assign these labels
for (var_name in names(nice_labels)) {
  var_label(gtys_analyse_2000_2021[[var_name]]) <- nice_labels[[var_name]]
}


gtys_analyse_2000_2021 <- gtys_analyse_2000_2021 %>% 
  select(Country, Year,curr_smk,Age,Sex,Grade,chew,smoking_adver,anti_media,smok_cigar_experimented,smokeless_chew_tried,smokeless_snuff_tried,smokeless_shisha_tried,smok_schl_mate,schl_tobacco_effects,tc_act,tc_smok_ban_minors,tc_smok_ban_public, tc_smok_ban_ads, tcbranded_item_owned,tobacco_from_friend)
```

#### Table 1

```
Table1=table1(~ Country +  Year + Age + Sex + Grade + chew + smoking_adver + anti_media + smok_cigar_experimented + smokeless_chew_tried + smokeless_snuff_tried + smokeless_shisha_tried + schl_tobacco_effects + tc_act + tc_smok_ban_minors + tc_smok_ban_public +  tc_smok_ban_ads +  tcbranded_item_owned,
       data = gtys_analyse_2000_2021)

knitr::kable(Table1, caption = "Summary Table 1 from GTYS")
```

Summary Table 1 from GTYS

|  | Overall |
| --- | --- |
|  | (N=439322) |
| Country |  |
| Algeria | 13714 (3.1%) |
| Angola | 1576 (0.4%) |
| Benin | 4329 (1.0%) |
| Botswana | 4127 (0.9%) |
| Burkina faso | 4447 (1.0%) |
| Burkina Faso | 6634 (1.5%) |
| Burundi | 2521 (0.6%) |
| Cameroon | 15599 (3.6%) |
| Cape Verde | 2019 (0.5%) |
| CAR | 2027 (0.5%) |
| Chad | 5333 (1.2%) |
| Comoros | 4620 (1.1%) |
| Congo | 3109 (0.7%) |
| DRC | 12717 (2.9%) |
| Equatorial Guinea | 3136 (0.7%) |
| Eritrea | 9639 (2.2%) |
| Ethiopia | 1868 (0.4%) |
| Gabon | 1781 (0.4%) |
| Gambia | 14544 (3.3%) |
| Ghana | 23949 (5.5%) |
| Guinea | 5038 (1.1%) |
| Ivory Coast | 10152 (2.3%) |
| Kenya | 17411 (4.0%) |
| Lesotho | 7573 (1.7%) |
| Liberia | 1739 (0.4%) |
| Madagascar | 4911 (1.1%) |
| Malawi | 7617 (1.7%) |
| Mali | 6227 (1.4%) |
| Mauritania | 15740 (3.6%) |
| Mauritius | 7812 (1.8%) |
| Mozambique | 13642 (3.1%) |
| Namibia | 8642 (2.0%) |
| Niger | 6193 (1.4%) |
| Nigeria | 5459 (1.2%) |
| Rwanda | 2284 (0.5%) |
| Senegal | 13208 (3.0%) |
| Seychelles | 5314 (1.2%) |
| Siera Leone | 2931 (0.7%) |
| Sierraleone | 6680 (1.5%) |
| South Africa | 28370 (6.5%) |
| STP | 8525 (1.9%) |
| Swaziland | 27211 (6.2%) |
| Tanzania | 16016 (3.6%) |
| Togo | 17879 (4.1%) |
| Uganda | 17452 (4.0%) |
| Zambia | 22263 (5.1%) |
| Zimbabwe | 15344 (3.5%) |
| Survey year |  |
| 2001 | 22626 (5.2%) |
| 2002 | 41637 (9.5%) |
| 2003 | 26756 (6.1%) |
| 2004 | 6231 (1.4%) |
| 2005 | 23145 (5.3%) |
| 2006 | 32930 (7.5%) |
| 2007 | 45376 (10.3%) |
| 2008 | 80144 (18.2%) |
| 2009 | 29087 (6.6%) |
| 2010 | 10101 (2.3%) |
| 2011 | 17660 (4.0%) |
| 2013 | 20748 (4.7%) |
| 2014 | 11130 (2.5%) |
| 2015 | 5295 (1.2%) |
| 2016 | 7981 (1.8%) |
| 2017 | 24929 (5.7%) |
| 2018 | 10118 (2.3%) |
| 2019 | 12609 (2.9%) |
| 2020 | 4320 (1.0%) |
| 2021 | 6499 (1.5%) |
| Age |  |
| 11 | 23059 (5.2%) |
| 12 | 37922 (8.6%) |
| 13 | 64182 (14.6%) |
| 14 | 85297 (19.4%) |
| 15 | 98981 (22.5%) |
| 16 | 68690 (15.6%) |
| 17 | 55456 (12.6%) |
| 18 | 5735 (1.3%) |
| Sex |  |
| Female | 220794 (50.3%) |
| Male | 218528 (49.7%) |
| School grade |  |
| 6 | 17423 (4.0%) |
| 7 | 58584 (13.3%) |
| 8 | 79185 (18.0%) |
| 9 | 100407 (22.9%) |
| 10 | 84468 (19.2%) |
| 11 | 54802 (12.5%) |
| 12 | 44453 (10.1%) |
| Used chewing tobacco |  |
| No | 424650 (96.7%) |
| Yes | 14672 (3.3%) |
| Saw smoking advertisement |  |
| No | 6092 (1.4%) |
| Yes | 433230 (98.6%) |
| Saw anti-smoking media |  |
| No | 93865 (21.4%) |
| Yes | 345457 (78.6%) |
| Ever tried cigars |  |
| No | 438451 (99.8%) |
| Yes | 871 (0.2%) |
| Ever tried smokeless chew |  |
| No | 439002 (99.9%) |
| Yes | 320 (0.1%) |
| Ever tried smokeless snuff |  |
| No | 438641 (99.8%) |
| Yes | 681 (0.2%) |
| Ever tried shisha |  |
| No | 437588 (99.6%) |
| Yes | 1734 (0.4%) |
| Taught about tobacco effects |  |
| No | 4737 (1.1%) |
| Yes | 434585 (98.9%) |
| Aware of tobacco control act |  |
| No | 439322 (100%) |
| Supports ban: sales to minors |  |
| No | 423951 (96.5%) |
| Yes | 15371 (3.5%) |
| Supports ban: smoking in public |  |
| No | 152362 (34.7%) |
| Yes | 286960 (65.3%) |
| Supports ban: tobacco advertising |  |
| No | 431901 (98.3%) |
| Yes | 7421 (1.7%) |
| Owns branded tobacco item |  |
| No | 370692 (84.4%) |
| Yes | 68630 (15.6%) |

Table 2: Prevalence of cigarette smoking

```
group_vars <- c(
  "Country", "Year", "Age", "Sex", "Grade",
  "chew", "smoking_adver", "anti_media",
  "smok_cigar_experimented", "smokeless_chew_tried", "smokeless_snuff_tried", "smokeless_shisha_tried",
  "schl_tobacco_effects", "tc_act",
  "tc_smok_ban_minors", "tc_smok_ban_public", "tc_smok_ban_ads", "tcbranded_item_owned"
)


library(dplyr)
library(tidyr)
library(purrr)
library(broom)
library(ggplot2)
library(forcats)

# Function to compute prevalence, CI and p-value
compute_stats <- function(data, var, outcome = "curr_smk") {
  data <- data %>% filter(!is.na(.data[[var]]), !is.na(.data[[outcome]]))
  
  summary <- data %>%
    group_by(.data[[var]]) %>%
    summarise(
      n = n(),
      cases = sum(.data[[outcome]]),
      prevalence = mean(.data[[outcome]]),
      .groups = "drop"
    ) %>%
    mutate(
      se = sqrt(prevalence * (1 - prevalence) / n),
      lower_95ci = prevalence - 1.96 * se,
      upper_95ci = prevalence + 1.96 * se,
      variable = var,
      level = paste0(var, ": ", .data[[var]])
    )
  
  chisq <- tryCatch({
    test <- chisq.test(table(data[[var]], data[[outcome]]))
    test$p.value
  }, error = function(e) NA)
  
  summary <- summary %>%
    mutate(p_value = chisq)
  
  return(summary)
}

# Run over all variables
results <- map_df(group_vars, ~ compute_stats(gtys_analyse_2000_2021, .x))

library(knitr)
library(kableExtra)
```

```
Attaching package: 'kableExtra'
```

```
The following object is masked from 'package:dplyr':

    group_rows
```

```
results %>%
  select(variable, level, n, cases, prevalence, lower_95ci, upper_95ci, p_value) %>%
  mutate(
    prevalence = round(prevalence * 100, 2),
    lower_95ci = round(lower_95ci * 100, 2),
    upper_95ci = round(upper_95ci * 100, 2),
    p_value = signif(p_value, 3)
  ) %>%
  kable(
    caption = "Prevalence of Current Smoking by Variable and Level (as %)",
    col.names = c("Variable", "Level", "N", "Cases", "Prev (%)", "Lower 95% CI (%)", "Upper 95% CI (%)", "P-Value"),
    align = "l"
  ) %>%
  kable_styling(bootstrap_options = c("striped", "hover", "condensed"), full_width = FALSE)
```

Prevalence of Current Smoking by Variable and Level (as %)

| Variable | Level | N | Cases | Prev (%) | Lower 95% CI (%) | Upper 95% CI (%) | P-Value |
| --- | --- | --- | --- | --- | --- | --- | --- |
| Country | Country: Algeria | 13714 | 823 | 6.00 | 5.60 | 6.40 | 0.0000000 |
| Country | Country: Angola | 1576 | 21 | 1.33 | 0.77 | 1.90 | 0.0000000 |
| Country | Country: Benin | 4329 | 446 | 10.30 | 9.40 | 11.21 | 0.0000000 |
| Country | Country: Botswana | 4127 | 473 | 11.46 | 10.49 | 12.43 | 0.0000000 |
| Country | Country: Burkina faso | 4447 | 4347 | 97.75 | 97.32 | 98.19 | 0.0000000 |
| Country | Country: Burkina Faso | 6634 | 716 | 10.79 | 10.05 | 11.54 | 0.0000000 |
| Country | Country: Burundi | 2521 | 132 | 5.24 | 4.37 | 6.11 | 0.0000000 |
| Country | Country: Cameroon | 15599 | 1015 | 6.51 | 6.12 | 6.89 | 0.0000000 |
| Country | Country: Cape Verde | 2019 | 107 | 5.30 | 4.32 | 6.28 | 0.0000000 |
| Country | Country: CAR | 2027 | 161 | 7.94 | 6.77 | 9.12 | 0.0000000 |
| Country | Country: Chad | 5333 | 341 | 6.39 | 5.74 | 7.05 | 0.0000000 |
| Country | Country: Comoros | 4620 | 239 | 5.17 | 4.53 | 5.81 | 0.0000000 |
| Country | Country: Congo | 3109 | 527 | 16.95 | 15.63 | 18.27 | 0.0000000 |
| Country | Country: DRC | 12717 | 886 | 6.97 | 6.52 | 7.41 | 0.0000000 |
| Country | Country: Equatorial Guinea | 3136 | 300 | 9.57 | 8.54 | 10.60 | 0.0000000 |
| Country | Country: Eritrea | 9639 | 192 | 1.99 | 1.71 | 2.27 | 0.0000000 |
| Country | Country: Ethiopia | 1868 | 54 | 2.89 | 2.13 | 3.65 | 0.0000000 |
| Country | Country: Gabon | 1781 | 154 | 8.65 | 7.34 | 9.95 | 0.0000000 |
| Country | Country: Gambia | 14544 | 1063 | 7.31 | 6.89 | 7.73 | 0.0000000 |
| Country | Country: Ghana | 23949 | 1001 | 4.18 | 3.93 | 4.43 | 0.0000000 |
| Country | Country: Guinea | 5038 | 435 | 8.63 | 7.86 | 9.41 | 0.0000000 |
| Country | Country: Ivory Coast | 10152 | 1482 | 14.60 | 13.91 | 15.28 | 0.0000000 |
| Country | Country: Kenya | 17411 | 1250 | 7.18 | 6.80 | 7.56 | 0.0000000 |
| Country | Country: Lesotho | 7573 | 980 | 12.94 | 12.18 | 13.70 | 0.0000000 |
| Country | Country: Liberia | 1739 | 42 | 2.42 | 1.69 | 3.14 | 0.0000000 |
| Country | Country: Madagascar | 4911 | 789 | 16.07 | 15.04 | 17.09 | 0.0000000 |
| Country | Country: Malawi | 7617 | 225 | 2.95 | 2.57 | 3.33 | 0.0000000 |
| Country | Country: Mali | 6227 | 1083 | 17.39 | 16.45 | 18.33 | 0.0000000 |
| Country | Country: Mauritania | 15740 | 2548 | 16.19 | 15.61 | 16.76 | 0.0000000 |
| Country | Country: Mauritius | 7812 | 1019 | 13.04 | 12.30 | 13.79 | 0.0000000 |
| Country | Country: Mozambique | 13642 | 367 | 2.69 | 2.42 | 2.96 | 0.0000000 |
| Country | Country: Namibia | 8642 | 1300 | 15.04 | 14.29 | 15.80 | 0.0000000 |
| Country | Country: Niger | 6193 | 705 | 11.38 | 10.59 | 12.17 | 0.0000000 |
| Country | Country: Nigeria | 5459 | 244 | 4.47 | 3.92 | 5.02 | 0.0000000 |
| Country | Country: Rwanda | 2284 | 63 | 2.76 | 2.09 | 3.43 | 0.0000000 |
| Country | Country: Senegal | 13208 | 1235 | 9.35 | 8.85 | 9.85 | 0.0000000 |
| Country | Country: Seychelles | 5314 | 925 | 17.41 | 16.39 | 18.43 | 0.0000000 |
| Country | Country: Siera Leone | 2931 | 232 | 7.92 | 6.94 | 8.89 | 0.0000000 |
| Country | Country: Sierraleone | 6680 | 225 | 3.37 | 2.94 | 3.80 | 0.0000000 |
| Country | Country: South Africa | 28370 | 4949 | 17.44 | 17.00 | 17.89 | 0.0000000 |
| Country | Country: STP | 8525 | 530 | 6.22 | 5.70 | 6.73 | 0.0000000 |
| Country | Country: Swaziland | 27211 | 2345 | 8.62 | 8.28 | 8.95 | 0.0000000 |
| Country | Country: Tanzania | 16016 | 373 | 2.33 | 2.10 | 2.56 | 0.0000000 |
| Country | Country: Togo | 17879 | 1169 | 6.54 | 6.18 | 6.90 | 0.0000000 |
| Country | Country: Uganda | 17452 | 1465 | 8.39 | 7.98 | 8.81 | 0.0000000 |
| Country | Country: Zambia | 22263 | 3234 | 14.53 | 14.06 | 14.99 | 0.0000000 |
| Country | Country: Zimbabwe | 15344 | 919 | 5.99 | 5.61 | 6.36 | 0.0000000 |
| Year | Year: 2001 | 22626 | 3177 | 14.04 | 13.59 | 14.49 | 0.0000000 |
| Year | Year: 2002 | 41637 | 5622 | 13.50 | 13.17 | 13.83 | 0.0000000 |
| Year | Year: 2003 | 26756 | 2290 | 8.56 | 8.22 | 8.89 | 0.0000000 |
| Year | Year: 2004 | 6231 | 1060 | 17.01 | 16.08 | 17.94 | 0.0000000 |
| Year | Year: 2005 | 23145 | 1691 | 7.31 | 6.97 | 7.64 | 0.0000000 |
| Year | Year: 2006 | 32930 | 6404 | 19.45 | 19.02 | 19.87 | 0.0000000 |
| Year | Year: 2007 | 45376 | 4184 | 9.22 | 8.95 | 9.49 | 0.0000000 |
| Year | Year: 2008 | 80144 | 7030 | 8.77 | 8.58 | 8.97 | 0.0000000 |
| Year | Year: 2009 | 29087 | 2487 | 8.55 | 8.23 | 8.87 | 0.0000000 |
| Year | Year: 2010 | 10101 | 551 | 5.45 | 5.01 | 5.90 | 0.0000000 |
| Year | Year: 2011 | 17660 | 2971 | 16.82 | 16.27 | 17.38 | 0.0000000 |
| Year | Year: 2013 | 20748 | 623 | 3.00 | 2.77 | 3.23 | 0.0000000 |
| Year | Year: 2014 | 11130 | 828 | 7.44 | 6.95 | 7.93 | 0.0000000 |
| Year | Year: 2015 | 5295 | 346 | 6.53 | 5.87 | 7.20 | 0.0000000 |
| Year | Year: 2016 | 7981 | 598 | 7.49 | 6.92 | 8.07 | 0.0000000 |
| Year | Year: 2017 | 24929 | 1270 | 5.09 | 4.82 | 5.37 | 0.0000000 |
| Year | Year: 2018 | 10118 | 938 | 9.27 | 8.71 | 9.84 | 0.0000000 |
| Year | Year: 2019 | 12609 | 456 | 3.62 | 3.29 | 3.94 | 0.0000000 |
| Year | Year: 2020 | 4320 | 143 | 3.31 | 2.78 | 3.84 | 0.0000000 |
| Year | Year: 2021 | 6499 | 462 | 7.11 | 6.48 | 7.73 | 0.0000000 |
| Age | Age: 11 | 23059 | 2406 | 10.43 | 10.04 | 10.83 | 0.0000000 |
| Age | Age: 12 | 37922 | 3261 | 8.60 | 8.32 | 8.88 | 0.0000000 |
| Age | Age: 13 | 64182 | 4678 | 7.29 | 7.09 | 7.49 | 0.0000000 |
| Age | Age: 14 | 85297 | 6833 | 8.01 | 7.83 | 8.19 | 0.0000000 |
| Age | Age: 15 | 98981 | 9208 | 9.30 | 9.12 | 9.48 | 0.0000000 |
| Age | Age: 16 | 68690 | 7858 | 11.44 | 11.20 | 11.68 | 0.0000000 |
| Age | Age: 17 | 55456 | 7755 | 13.98 | 13.70 | 14.27 | 0.0000000 |
| Age | Age: 18 | 5735 | 1132 | 19.74 | 18.71 | 20.77 | 0.0000000 |
| Sex | Sex: Female | 220794 | 15320 | 6.94 | 6.83 | 7.04 | 0.0000000 |
| Sex | Sex: Male | 218528 | 27811 | 12.73 | 12.59 | 12.87 | 0.0000000 |
| Grade | Grade: 6 | 17423 | 1659 | 9.52 | 9.09 | 9.96 | 0.0000000 |
| Grade | Grade: 7 | 58584 | 4561 | 7.79 | 7.57 | 8.00 | 0.0000000 |
| Grade | Grade: 8 | 79185 | 7646 | 9.66 | 9.45 | 9.86 | 0.0000000 |
| Grade | Grade: 9 | 100407 | 8744 | 8.71 | 8.53 | 8.88 | 0.0000000 |
| Grade | Grade: 10 | 84468 | 8598 | 10.18 | 9.98 | 10.38 | 0.0000000 |
| Grade | Grade: 11 | 54802 | 6462 | 11.79 | 11.52 | 12.06 | 0.0000000 |
| Grade | Grade: 12 | 44453 | 5461 | 12.28 | 11.98 | 12.59 | 0.0000000 |
| chew | chew: No | 424650 | 39822 | 9.38 | 9.29 | 9.47 | 0.0000000 |
| chew | chew: Yes | 14672 | 3309 | 22.55 | 21.88 | 23.23 | 0.0000000 |
| smoking\_adver | smoking\_adver: No | 6092 | 368 | 6.04 | 5.44 | 6.64 | 0.0000000 |
| smoking\_adver | smoking\_adver: Yes | 433230 | 42763 | 9.87 | 9.78 | 9.96 | 0.0000000 |
| anti\_media | anti\_media: No | 93865 | 5516 | 5.88 | 5.73 | 6.03 | 0.0000000 |
| anti\_media | anti\_media: Yes | 345457 | 37615 | 10.89 | 10.78 | 10.99 | 0.0000000 |
| smok\_cigar\_experimented | smok\_cigar\_experimented: No | 438451 | 43008 | 9.81 | 9.72 | 9.90 | 0.0000248 |
| smok\_cigar\_experimented | smok\_cigar\_experimented: Yes | 871 | 123 | 14.12 | 11.81 | 16.43 | 0.0000248 |
| smokeless\_chew\_tried | smokeless\_chew\_tried: No | 439002 | 43023 | 9.80 | 9.71 | 9.89 | 0.0000000 |
| smokeless\_chew\_tried | smokeless\_chew\_tried: Yes | 320 | 108 | 33.75 | 28.57 | 38.93 | 0.0000000 |
| smokeless\_snuff\_tried | smokeless\_snuff\_tried: No | 438641 | 43013 | 9.81 | 9.72 | 9.89 | 0.0000000 |
| smokeless\_snuff\_tried | smokeless\_snuff\_tried: Yes | 681 | 118 | 17.33 | 14.48 | 20.17 | 0.0000000 |
| smokeless\_shisha\_tried | smokeless\_shisha\_tried: No | 437588 | 42791 | 9.78 | 9.69 | 9.87 | 0.0000000 |
| smokeless\_shisha\_tried | smokeless\_shisha\_tried: Yes | 1734 | 340 | 19.61 | 17.74 | 21.48 | 0.0000000 |
| schl\_tobacco\_effects | schl\_tobacco\_effects: No | 4737 | 486 | 10.26 | 9.40 | 11.12 | 0.3160000 |
| schl\_tobacco\_effects | schl\_tobacco\_effects: Yes | 434585 | 42645 | 9.81 | 9.72 | 9.90 | 0.3160000 |
| tc\_act | tc\_act: No | 439322 | 43131 | 9.82 | 9.73 | 9.91 | 0.0000000 |
| tc\_smok\_ban\_minors | tc\_smok\_ban\_minors: No | 423951 | 42320 | 9.98 | 9.89 | 10.07 | 0.0000000 |
| tc\_smok\_ban\_minors | tc\_smok\_ban\_minors: Yes | 15371 | 811 | 5.28 | 4.92 | 5.63 | 0.0000000 |
| tc\_smok\_ban\_public | tc\_smok\_ban\_public: No | 152362 | 16793 | 11.02 | 10.86 | 11.18 | 0.0000000 |
| tc\_smok\_ban\_public | tc\_smok\_ban\_public: Yes | 286960 | 26338 | 9.18 | 9.07 | 9.28 | 0.0000000 |
| tc\_smok\_ban\_ads | tc\_smok\_ban\_ads: No | 431901 | 42916 | 9.94 | 9.85 | 10.03 | 0.0000000 |
| tc\_smok\_ban\_ads | tc\_smok\_ban\_ads: Yes | 7421 | 215 | 2.90 | 2.52 | 3.28 | 0.0000000 |
| tcbranded\_item\_owned | tcbranded\_item\_owned: No | 370692 | 32323 | 8.72 | 8.63 | 8.81 | 0.0000000 |
| tcbranded\_item\_owned | tcbranded\_item\_owned: Yes | 68630 | 10808 | 15.75 | 15.48 | 16.02 | 0.0000000 |

Table 4: Bivariate and multivariate logistic regression of harmful alcohol uptake

```
# Load libraries
library(dplyr)
library(purrr)
library(broom)
library(MASS)
```

```
Attaching package: 'MASS'
```

```
The following object is masked from 'package:dplyr':

    select
```

```
library(knitr)
library(kableExtra)

# Grouping variables
group_vars <- c(
  "Country", "Year", "Age", "Sex", "Grade",
  "chew", "smoking_adver", "anti_media",
  "smok_cigar_experimented", "smokeless_chew_tried", 
  "smokeless_snuff_tried", "smokeless_shisha_tried", "schl_tobacco_effects",
  "tc_smok_ban_minors", "tc_smok_ban_public", 
  "tc_smok_ban_ads", "tcbranded_item_owned"
)

# Outcome variable is "curr_smk" (coded 0/1 already)
# If not, make sure it is binary
# gtys_analyse_2000_2021 <- gtys_analyse_2000_2021 %>%
#   mutate(curr_smk = ifelse(curr_smk == "Yes", 1, 0))

# Safe GLM function
safe_model <- safely(function(var) {
  fml <- as.formula(paste("curr_smk ~", var))
  model <- glm(fml, 
               data = gtys_analyse_2000_2021, 
               family = binomial, 
               na.action = na.omit)
  broom::tidy(model, exponentiate = TRUE, conf.int = TRUE) %>%
    filter(term != "(Intercept)") %>%
    mutate(variable = var, model = "Crude") %>%
    dplyr::select(variable, term, estimate, conf.low, conf.high, p.value, model)
})

# Run crude models
uni_results_list <- map(group_vars, safe_model)
uni_results <- uni_results_list %>%
  map("result") %>%
  compact() %>%
  bind_rows()

# Adjusted model
full_formula <- as.formula(
  paste("curr_smk ~", paste(group_vars, collapse = " + "))
)

valid_vars <- map_lgl(group_vars, function(v) {
  levels_count <- gtys_analyse_2000_2021 %>%
    filter(!is.na(.data[[v]])) %>%
    distinct(.data[[v]]) %>%
    nrow()
  levels_count > 1
})

# Keep only variables with at least 2 levels
group_vars_ok <- group_vars[valid_vars]
group_vars_ok
```

```
 [1] "Country"                 "Year"                   
 [3] "Age"                     "Sex"                    
 [5] "Grade"                   "chew"                   
 [7] "smoking_adver"           "anti_media"             
 [9] "smok_cigar_experimented" "smokeless_chew_tried"   
[11] "smokeless_snuff_tried"   "smokeless_shisha_tried" 
[13] "schl_tobacco_effects"    "tc_smok_ban_minors"     
[15] "tc_smok_ban_public"      "tc_smok_ban_ads"        
[17] "tcbranded_item_owned"
```

```
full_formula <- as.formula(
  paste("curr_smk ~", paste(group_vars_ok, collapse = " + "))
)
full_model <- glm(
  full_formula,
  data = gtys_analyse_2000_2021,
  family = binomial,
  na.action = na.omit,
  control = glm.control(maxit = 100, epsilon = 1e-8)
)

# Stepwise selection
step_model <- stepAIC(full_model, trace = FALSE, direction = "both")

# Tidy adjusted results
multi_results <- broom::tidy(step_model, exponentiate = TRUE, conf.int = TRUE) %>%
  filter(term != "(Intercept)") %>%
  mutate(
    variable = gsub("^(.*?)\\b.*$", "\\1", term),
    model    = "Adjusted"
  ) %>%
  dplyr::select(variable, term, estimate, conf.low, conf.high, p.value, model)

# Combine
results_table <- bind_rows(uni_results, multi_results) %>%
  mutate(
    term = gsub(paste0("^", variable), "", term),
    term = ifelse(term == "", "(reference)", term)
  ) %>%
  arrange(variable, model, term) %>%
  rename(
    `Odds Ratio`   = estimate,
    `95% CI Lower` = conf.low,
    `95% CI Upper` = conf.high,
    `p-value`      = p.value
  )
```

```
Warning: There was 1 warning in `mutate()`.
ℹ In argument: `term = gsub(paste0("^", variable), "", term)`.
Caused by warning in `gsub()`:
! argument 'pattern' has length > 1 and only the first element will be used
```

```
# Display table
results_table %>%
  kable(
    caption = "Crude and Adjusted Odds Ratios for Current Smoking",
    col.names = c("Variable", "Level", "Odds Ratio", "95% CI Lower", "95% CI Upper", "p-value", "Model"),
    digits = c(0, 0, 2, 2, 2, 3, NA),
    format = "html"
  ) %>%
  kable_styling(full_width = FALSE, position = "center")
```

Crude and Adjusted Odds Ratios for Current Smoking

| Variable | Level | Odds Ratio | 95% CI Lower | 95% CI Upper | p-value | Model |
| --- | --- | --- | --- | --- | --- | --- |
|  | Age12 | 0.93 | 0.86 | 1.00 | 0.066 | Adjusted |
|  | Age13 | 0.73 | 0.67 | 0.79 | 0.000 | Adjusted |
|  | Age14 | 0.89 | 0.83 | 0.96 | 0.002 | Adjusted |
|  | Age15 | 1.04 | 0.96 | 1.12 | 0.340 | Adjusted |
|  | Age16 | 1.22 | 1.13 | 1.32 | 0.000 | Adjusted |
|  | Age17 | 1.67 | 1.54 | 1.80 | 0.000 | Adjusted |
|  | Age18 | 1.45 | 1.31 | 1.61 | 0.000 | Adjusted |
|  | Angola | 0.08 | 0.05 | 0.12 | 0.000 | Adjusted |
|  | Benin | 0.70 | 0.59 | 0.82 | 0.000 | Adjusted |
|  | Botswana | 1.27 | 1.11 | 1.45 | 0.001 | Adjusted |
|  | Burkina Faso | 0.84 | 0.73 | 0.96 | 0.010 | Adjusted |
|  | Burkina faso | 380.55 | 301.05 | 485.33 | 0.000 | Adjusted |
|  | Burundi | 0.51 | 0.42 | 0.63 | 0.000 | Adjusted |
|  | CAR | 0.75 | 0.62 | 0.91 | 0.004 | Adjusted |
|  | Cameroon | 0.66 | 0.58 | 0.75 | 0.000 | Adjusted |
|  | Cape Verde | 0.63 | 0.50 | 0.77 | 0.000 | Adjusted |
|  | Chad | 0.80 | 0.69 | 0.94 | 0.005 | Adjusted |
|  | Comoros | 1.11 | 0.94 | 1.30 | 0.213 | Adjusted |
|  | Congo | 0.87 | 0.73 | 1.03 | 0.101 | Adjusted |
|  | DRC | 0.89 | 0.78 | 1.01 | 0.077 | Adjusted |
|  | Equatorial Guinea | 1.09 | 0.93 | 1.28 | 0.275 | Adjusted |
|  | Eritrea | 0.13 | 0.10 | 0.15 | 0.000 | Adjusted |
|  | Ethiopia | 0.23 | 0.17 | 0.30 | 0.000 | Adjusted |
|  | Gabon | 0.57 | 0.46 | 0.72 | 0.000 | Adjusted |
|  | Gambia | 0.93 | 0.79 | 1.10 | 0.399 | Adjusted |
|  | Ghana | 0.32 | 0.28 | 0.37 | 0.000 | Adjusted |
|  | Grade10 | 0.84 | 0.76 | 0.92 | 0.000 | Adjusted |
|  | Grade11 | 0.82 | 0.75 | 0.90 | 0.000 | Adjusted |
|  | Grade12 | 0.76 | 0.69 | 0.84 | 0.000 | Adjusted |
|  | Grade7 | 0.85 | 0.78 | 0.93 | 0.001 | Adjusted |
|  | Grade8 | 0.99 | 0.91 | 1.08 | 0.843 | Adjusted |
|  | Grade9 | 0.82 | 0.75 | 0.89 | 0.000 | Adjusted |
|  | Guinea | 0.81 | 0.70 | 0.93 | 0.004 | Adjusted |
|  | Ivory Coast | 1.45 | 1.27 | 1.66 | 0.000 | Adjusted |
|  | Kenya | 0.73 | 0.66 | 0.80 | 0.000 | Adjusted |
|  | Lesotho | 1.30 | 1.16 | 1.46 | 0.000 | Adjusted |
|  | Liberia | 0.22 | 0.16 | 0.31 | 0.000 | Adjusted |
|  | Madagascar | 1.61 | 1.41 | 1.85 | 0.000 | Adjusted |
|  | Malawi | 0.23 | 0.19 | 0.28 | 0.000 | Adjusted |
|  | Mali | 1.62 | 1.43 | 1.83 | 0.000 | Adjusted |
|  | Mauritania | 1.31 | 1.16 | 1.47 | 0.000 | Adjusted |
|  | Mauritius | 1.62 | 1.40 | 1.87 | 0.000 | Adjusted |
|  | Mozambique | 0.35 | 0.31 | 0.40 | 0.000 | Adjusted |
|  | Namibia | 1.09 | 0.92 | 1.29 | 0.336 | Adjusted |
|  | Niger | 0.88 | 0.76 | 1.01 | 0.059 | Adjusted |
|  | Nigeria | 0.43 | 0.36 | 0.50 | 0.000 | Adjusted |
|  | Rwanda | 0.24 | 0.18 | 0.31 | 0.000 | Adjusted |
|  | STP | 0.39 | 0.34 | 0.45 | 0.000 | Adjusted |
|  | Senegal | 1.29 | 1.16 | 1.43 | 0.000 | Adjusted |
|  | SexMale | 2.05 | 2.00 | 2.09 | 0.000 | Adjusted |
|  | Seychelles | 3.05 | 2.71 | 3.44 | 0.000 | Adjusted |
|  | Siera Leone | 0.74 | 0.62 | 0.87 | 0.000 | Adjusted |
|  | Sierraleone | 0.40 | 0.32 | 0.49 | 0.000 | Adjusted |
|  | South Africa | 1.62 | 1.47 | 1.79 | 0.000 | Adjusted |
|  | Swaziland | 0.55 | 0.49 | 0.63 | 0.000 | Adjusted |
|  | Tanzania | 0.18 | 0.15 | 0.21 | 0.000 | Adjusted |
|  | Togo | 0.78 | 0.70 | 0.86 | 0.000 | Adjusted |
|  | Uganda | 0.80 | 0.72 | 0.89 | 0.000 | Adjusted |
|  | Year2002 | 0.70 | 0.65 | 0.76 | 0.000 | Adjusted |
|  | Year2003 | 0.89 | 0.81 | 0.99 | 0.030 | Adjusted |
|  | Year2004 | 1.18 | 0.99 | 1.39 | 0.061 | Adjusted |
|  | Year2005 | 0.95 | 0.87 | 1.04 | 0.282 | Adjusted |
|  | Year2006 | 0.91 | 0.83 | 0.99 | 0.025 | Adjusted |
|  | Year2007 | 0.64 | 0.59 | 0.69 | 0.000 | Adjusted |
|  | Year2008 | 0.65 | 0.60 | 0.70 | 0.000 | Adjusted |
|  | Year2009 | 0.75 | 0.70 | 0.80 | 0.000 | Adjusted |
|  | Year2010 | NA | NA | NA | NA | Adjusted |
|  | Year2011 | 0.72 | 0.66 | 0.78 | 0.000 | Adjusted |
|  | Year2013 | 0.14 | 0.12 | 0.17 | 0.000 | Adjusted |
|  | Year2014 | 0.52 | 0.45 | 0.61 | 0.000 | Adjusted |
|  | Year2015 | 0.12 | 0.10 | 0.14 | 0.000 | Adjusted |
|  | Year2016 | 0.58 | 0.49 | 0.67 | 0.000 | Adjusted |
|  | Year2017 | 0.42 | 0.36 | 0.50 | 0.000 | Adjusted |
|  | Year2018 | 0.41 | 0.36 | 0.47 | 0.000 | Adjusted |
|  | Year2019 | 0.20 | 0.17 | 0.23 | 0.000 | Adjusted |
|  | Year2020 | 0.13 | 0.11 | 0.17 | 0.000 | Adjusted |
|  | Year2021 | 0.19 | 0.16 | 0.22 | 0.000 | Adjusted |
|  | Zambia | 1.73 | 1.57 | 1.90 | 0.000 | Adjusted |
|  | Zimbabwe | 0.49 | 0.42 | 0.56 | 0.000 | Adjusted |
|  | anti\_mediaYes | 0.59 | 0.54 | 0.64 | 0.000 | Adjusted |
|  | chewYes | 2.11 | 2.00 | 2.21 | 0.000 | Adjusted |
|  | schl\_tobacco\_effectsYes | 0.79 | 0.69 | 0.89 | 0.000 | Adjusted |
|  | smokeless\_chew\_triedYes | 1.68 | 1.29 | 2.17 | 0.000 | Adjusted |
|  | smokeless\_shisha\_triedYes | 3.89 | 3.36 | 4.50 | 0.000 | Adjusted |
|  | smokeless\_snuff\_triedYes | 0.46 | 0.37 | 0.57 | 0.000 | Adjusted |
|  | smoking\_adverYes | 1.25 | 1.10 | 1.42 | 0.001 | Adjusted |
|  | tc\_smok\_ban\_minorsYes | 0.89 | 0.79 | 1.01 | 0.063 | Adjusted |
|  | tc\_smok\_ban\_publicYes | 0.79 | 0.77 | 0.81 | 0.000 | Adjusted |
|  | tcbranded\_item\_ownedYes | 1.85 | 1.80 | 1.90 | 0.000 | Adjusted |
| Age | Age12 | 0.81 | 0.76 | 0.85 | 0.000 | Crude |
| Age | Age13 | 0.67 | 0.64 | 0.71 | 0.000 | Crude |
| Age | Age14 | 0.75 | 0.71 | 0.79 | 0.000 | Crude |
| Age | Age15 | 0.88 | 0.84 | 0.92 | 0.000 | Crude |
| Age | Age16 | 1.11 | 1.06 | 1.16 | 0.000 | Crude |
| Age | Age17 | 1.40 | 1.33 | 1.47 | 0.000 | Crude |
| Age | Age18 | 2.11 | 1.95 | 2.28 | 0.000 | Crude |
| Country | Angola | 0.21 | 0.13 | 0.32 | 0.000 | Crude |
| Country | Benin | 1.80 | 1.59 | 2.03 | 0.000 | Crude |
| Country | Botswana | 2.03 | 1.80 | 2.28 | 0.000 | Crude |
| Country | Burkina Faso | 1.90 | 1.71 | 2.10 | 0.000 | Crude |
| Country | Burkina faso | 680.89 | 554.71 | 845.26 | 0.000 | Crude |
| Country | Burundi | 0.87 | 0.71 | 1.04 | 0.134 | Crude |
| Country | CAR | 1.35 | 1.13 | 1.61 | 0.001 | Crude |
| Country | Cameroon | 1.09 | 0.99 | 1.20 | 0.075 | Crude |
| Country | Cape Verde | 0.88 | 0.71 | 1.07 | 0.212 | Crude |
| Country | Chad | 1.07 | 0.94 | 1.22 | 0.309 | Crude |
| Country | Comoros | 0.85 | 0.74 | 0.99 | 0.037 | Crude |
| Country | Congo | 3.20 | 2.84 | 3.59 | 0.000 | Crude |
| Country | DRC | 1.17 | 1.06 | 1.29 | 0.001 | Crude |
| Country | Equatorial Guinea | 1.66 | 1.44 | 1.90 | 0.000 | Crude |
| Country | Eritrea | 0.32 | 0.27 | 0.37 | 0.000 | Crude |
| Country | Ethiopia | 0.47 | 0.35 | 0.61 | 0.000 | Crude |
| Country | Gabon | 1.48 | 1.23 | 1.77 | 0.000 | Crude |
| Country | Gambia | 1.24 | 1.12 | 1.36 | 0.000 | Crude |
| Country | Ghana | 0.68 | 0.62 | 0.75 | 0.000 | Crude |
| Country | Guinea | 1.48 | 1.31 | 1.67 | 0.000 | Crude |
| Country | Ivory Coast | 2.68 | 2.45 | 2.93 | 0.000 | Crude |
| Country | Kenya | 1.21 | 1.11 | 1.33 | 0.000 | Crude |
| Country | Lesotho | 2.33 | 2.11 | 2.57 | 0.000 | Crude |
| Country | Liberia | 0.39 | 0.28 | 0.52 | 0.000 | Crude |
| Country | Madagascar | 3.00 | 2.70 | 3.33 | 0.000 | Crude |
| Country | Malawi | 0.48 | 0.41 | 0.55 | 0.000 | Crude |
| Country | Mali | 3.30 | 3.00 | 3.63 | 0.000 | Crude |
| Country | Mauritania | 3.03 | 2.79 | 3.29 | 0.000 | Crude |
| Country | Mauritius | 2.35 | 2.13 | 2.59 | 0.000 | Crude |
| Country | Mozambique | 0.43 | 0.38 | 0.49 | 0.000 | Crude |
| Country | Namibia | 2.77 | 2.53 | 3.04 | 0.000 | Crude |
| Country | Niger | 2.01 | 1.81 | 2.24 | 0.000 | Crude |
| Country | Nigeria | 0.73 | 0.63 | 0.85 | 0.000 | Crude |
| Country | Rwanda | 0.44 | 0.34 | 0.57 | 0.000 | Crude |
| Country | STP | 1.04 | 0.93 | 1.16 | 0.513 | Crude |
| Country | Senegal | 1.62 | 1.47 | 1.77 | 0.000 | Crude |
| Country | Seychelles | 3.30 | 2.99 | 3.65 | 0.000 | Crude |
| Country | Siera Leone | 1.35 | 1.15 | 1.56 | 0.000 | Crude |
| Country | Sierraleone | 0.55 | 0.47 | 0.63 | 0.000 | Crude |
| Country | South Africa | 3.31 | 3.07 | 3.58 | 0.000 | Crude |
| Country | Swaziland | 1.48 | 1.36 | 1.60 | 0.000 | Crude |
| Country | Tanzania | 0.37 | 0.33 | 0.42 | 0.000 | Crude |
| Country | Togo | 1.10 | 1.00 | 1.20 | 0.052 | Crude |
| Country | Uganda | 1.44 | 1.31 | 1.57 | 0.000 | Crude |
| Country | Zambia | 2.66 | 2.46 | 2.88 | 0.000 | Crude |
| Country | Zimbabwe | 1.00 | 0.91 | 1.10 | 0.966 | Crude |
| Grade | Grade10 | 1.08 | 1.02 | 1.14 | 0.009 | Crude |
| Grade | Grade11 | 1.27 | 1.20 | 1.34 | 0.000 | Crude |
| Grade | Grade12 | 1.33 | 1.26 | 1.41 | 0.000 | Crude |
| Grade | Grade7 | 0.80 | 0.76 | 0.85 | 0.000 | Crude |
| Grade | Grade8 | 1.02 | 0.96 | 1.07 | 0.587 | Crude |
| Grade | Grade9 | 0.91 | 0.86 | 0.96 | 0.000 | Crude |
| Sex | SexMale | 1.96 | 1.92 | 2.00 | 0.000 | Crude |
| Year | Year2002 | 0.96 | 0.91 | 1.00 | 0.058 | Crude |
| Year | Year2003 | 0.57 | 0.54 | 0.61 | 0.000 | Crude |
| Year | Year2004 | 1.25 | 1.16 | 1.35 | 0.000 | Crude |
| Year | Year2005 | 0.48 | 0.45 | 0.51 | 0.000 | Crude |
| Year | Year2006 | 1.48 | 1.41 | 1.55 | 0.000 | Crude |
| Year | Year2007 | 0.62 | 0.59 | 0.65 | 0.000 | Crude |
| Year | Year2008 | 0.59 | 0.56 | 0.62 | 0.000 | Crude |
| Year | Year2009 | 0.57 | 0.54 | 0.61 | 0.000 | Crude |
| Year | Year2010 | 0.35 | 0.32 | 0.39 | 0.000 | Crude |
| Year | Year2011 | 1.24 | 1.17 | 1.31 | 0.000 | Crude |
| Year | Year2013 | 0.19 | 0.17 | 0.21 | 0.000 | Crude |
| Year | Year2014 | 0.49 | 0.45 | 0.53 | 0.000 | Crude |
| Year | Year2015 | 0.43 | 0.38 | 0.48 | 0.000 | Crude |
| Year | Year2016 | 0.50 | 0.45 | 0.54 | 0.000 | Crude |
| Year | Year2017 | 0.33 | 0.31 | 0.35 | 0.000 | Crude |
| Year | Year2018 | 0.63 | 0.58 | 0.68 | 0.000 | Crude |
| Year | Year2019 | 0.23 | 0.21 | 0.25 | 0.000 | Crude |
| Year | Year2020 | 0.21 | 0.18 | 0.25 | 0.000 | Crude |
| Year | Year2021 | 0.47 | 0.42 | 0.52 | 0.000 | Crude |
| anti\_media | anti\_mediaYes | 1.96 | 1.90 | 2.02 | 0.000 | Crude |
| chew | chewYes | 2.81 | 2.70 | 2.93 | 0.000 | Crude |
| schl\_tobacco\_effects | schl\_tobacco\_effectsYes | 0.95 | 0.87 | 1.05 | 0.304 | Crude |
| smok\_cigar\_experimented | smok\_cigar\_experimentedYes | 1.51 | 1.24 | 1.82 | 0.000 | Crude |
| smokeless\_chew\_tried | smokeless\_chew\_triedYes | 4.69 | 3.71 | 5.90 | 0.000 | Crude |
| smokeless\_shisha\_tried | smokeless\_shisha\_triedYes | 2.25 | 1.99 | 2.53 | 0.000 | Crude |
| smokeless\_snuff\_tried | smokeless\_snuff\_triedYes | 1.93 | 1.57 | 2.34 | 0.000 | Crude |
| smoking\_adver | smoking\_adverYes | 1.70 | 1.53 | 1.90 | 0.000 | Crude |
| tc\_smok\_ban\_ads | tc\_smok\_ban\_adsYes | 0.27 | 0.24 | 0.31 | 0.000 | Crude |
| tc\_smok\_ban\_minors | tc\_smok\_ban\_minorsYes | 0.50 | 0.47 | 0.54 | 0.000 | Crude |
| tc\_smok\_ban\_public | tc\_smok\_ban\_publicYes | 0.82 | 0.80 | 0.83 | 0.000 | Crude |
| tcbranded\_item\_owned | tcbranded\_item\_ownedYes | 1.96 | 1.91 | 2.00 | 0.000 | Crude |
